# Verifying Infectious Disease Scenario Planning for Geographically Diverse Populations

**DOI:** 10.1101/2024.10.15.24314208

**Authors:** Jessica R. Conrad, Paul W. Fenimore, Kelly R. Moran, Marisa C. Eisenberg

**Author notes:** Corresponding author (J.R. Conrad). Co-senior author (Kelly R. Moran, Marisa C. Eisenberg). (P.W. Fenimore); (K.R. Moran); (M.C.Eisenberg).

## Abstract

In the face of the COVID-19 pandemic, the literature saw a spike in publications for epidemic models, and a renewed interest in capturing contact networks and geographic movement of populations. There remains a general lack of consensus in the modeling community around best practices for spatiotemporal epi-modeling, specifically as it pertains to the infection rate formulation and the underlying contact or mixing model.

In this work, we mathematically verify several common modeling assumptions in the literature, to prove when certain choices can provide consistent results across different geographic resolutions, population densities and patterns, and mixing assumptions. The most common infection rate formulation, a computationally low cost *per capita* infection rate assumption, fails the consistency tests for heterogeneous populations and non-symmetric mixing assumptions. The largest numerical errors occur in the limit of lowest symmetry, whether as sparse geography or preferential travel to highly-populated locations. Future modeling efforts in spatiotemporal disease modeling should be wary of this limitation, particularly when working with more heterogenous or less dense populations.

Our results provide guidance for testing that a model preserves desirable properties even when model inputs mask potential problems due to symmetry or homogeneity. We also provide a recipe for performing this type of validation with the objective of strengthening decision support tools.

**Highlights:**

- Define common modeling options from the literature for spatiotemporal epidemic models
- Verify common modeling assumptions are consistent for varying population densities and patterns, resolutions, and underlying mixing or contact assumptions
- Provide simulation examples of model misspecification and the resulting implications on scenario planning

## 1. Introduction

The number of published spatiotemporal epidemic models increased in 2020, in reaction to the global COVID-19 outbreak. In a recent survey of the literature, Pujante-Otalora, Canovas-Segura, Campos and Juarez (2023) found that most spatiotemporal models that focus on specific diseases are centered around respiratory-transmitted illnesses such as COVID-19, MERS, SARS-1, H1N1, and seasonal influenzas.

With the increased interest in the utility of these models for decision support, it is becoming increasingly important to have tools with which to *verify* and *validate* the models being proposed. One way to define verification versus validation is the following (Thacker, Doebling, Hemez, Anderson, Pepin and Rodriguez, 2004):

- **Verification:** “the process of determining that a model implementation accurately represents the developer’s conceptual description of the model and its solution; that is verifying model dynamics meet assumptions of the model;”
- **Validation:** “the process of determining the degree to which a model is an accurate representation of the real world from the perspective of the intended uses of the model; that is validating the model against data.”

Together these provide assurance that a model is sufficient for its intended use; in this context epidemic decision support. While many spatiotemporal epidemic models are validated against real or synthetic data (Pujante-Otalora et al., 2023), they are not necessarily verified. That is, while the model can be fit to the data, it remains to be seen that the underlying model assumptions are correct or match their intended purpose (Czocher, Stillman and Brown, 2018), particularly as model simplifications are often introduced, e.g. to improve computational efficiency or to make parameterization or analysis tractable. Without a verified model, there is a risk of inaccuracy for forseeable reasons. Furthermore, inference made on model parameters may lead to unsupported conclusions about underlying disease dynamics.

For spatiotemporal epidemic models, there are many possible choices for how to represent the spatial structure, ranging from those where nodes represent individuals to regular grids or politically-defined regions (Pujante-Otalora et al., 2023; Mills and Riley, 2014; Mourant, Fenimore, Manore and McMahon, 2018; Lee, Li, Liu and LeDuc, 2021; Long, Nohdurft and Spinler, 2018; Cao, Zhu, Wang, Tong, Tian, Dai and Ma, 2022; Yashima and Sasaki, 2014; Wardle, Bhatia, Kraemer, Nouvellet and Cori, 2023; LaBute, McMahon, Brown, Manore and Fair, 2014). Spatial epidemic models often trade tractability for resolution, leading to interactions with model assumptions—when is it appropriate to combine different population groups or individuals to form a single larger compartment? When would a compartmental model structured around political divisions (national, regional, county, census tract) be verifiable? When is it necessary to transition to regular geographical grids or a more individual-based model?

All of these questions also bring questions of computational tractability: highly spatially-resolved models are often compute- and memory-intensive, presenting barriers to rapid decision support. Decision support may be made in the absence of a verified model, which presents difficulties in making *robust inference*, for example with standarized assessment methodology (Koopman, 2004). The purpose of such evaluation is to establish how alternative assumptions around (in this case) spatial structure may impact model conclusions and inferences.

In this work, we restrict our ourselves to geographically-gridded models with differing assumptions about geographical spread of disease. We perform verification and inference robustness assessment for common spatio-temporal modeling choices found in the epidemiological modeling literature. Of particular interest here is the infection rate formulation and geographical mixing model, and the implications of these choices for model properites as a function of grid resolution. Because we find that in some cases unexpected behavior can arise, this can impact inferences about the time of an epidemic peak and the basic reproductive number, ℛ_0_.

## 2. Methods

### 2.1. General Model and Options

The following section outlines the modeling framework, modeling options, and evaluation metrics. Table 1 summarizes all variables as defined for the modeling framework and evaluation metrics. Table 2 provides a brief overview of the modeling options explored in this paper, which are described more in depth in Section 2.1.2 and Section 2.1.3.

**Table 1.** Summary of variable and parameter definitions for the mixing model, infection model, and evaluation metrics. It is generally assumed that only *β, μ*, and *s*_*d*_ are unknown.

| Variable | Definition |
| --- | --- |
| $M$ | $n \times n$ connectivity or mixing matrix; entries $m_{ij}$ are connectivity of cell $i$ to $j$ |
| $n$ | total number of cells in the population grid |
| $\kappa^*(r_{ij})$ | diffusion kernel given the distance $r_{ij}$; i.e. relative contact between cell $i$ and $j$ |
| $r_{ij}$ | center-to-center distance between cell $i$ and $j$ |
| $s_d$ | distance below which contact rates saturate; i.e. characteristic distance |
| $\Delta x$ | cell size of population grid; i.e. resolution |
| $\gamma$ | mixing parameter of $\kappa^T(r_{ij})$; $\gamma = -4$ |
| $N_i$ | total number of people in cell $i$ |
| $S_{i,t}$ | susceptible people in cell $i$ at time $t$ |
| $I_{i,t}$ | infected people in cell $i$ at time $t$ |
| $R_{i,t}$ | recovered people in cell $i$ at time $t$ |
| $\lambda_{i,t}^*$ | force of infection in cell $i$ at time $t$ |
| $\beta$ | rate of transmission per infectious contact |
| $\mu$ | average rate of recovery |
| $\mathcal{R}_0$ | basic reproduction number; dimensionless bifurcation parameter of infection model |

**Table 2.**
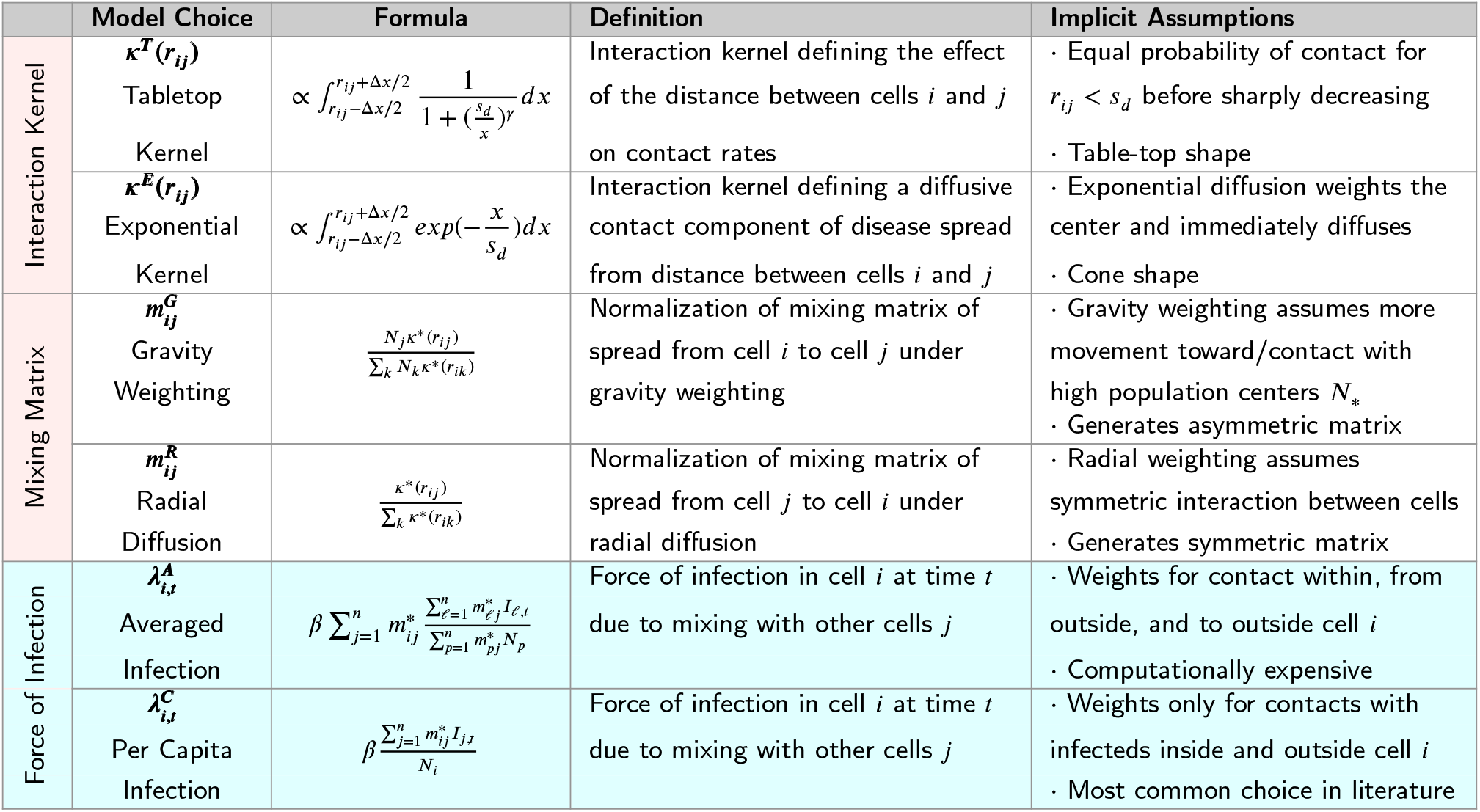
Summary of different model choices that this paper explores in depth. Two options are available for each choice of interaction kernel, mixing matrix normalization, and infection rate (force of infection). These model choices can be seen throughout the literature (Wu, Riley and Leung, 2007; Mills and Riley, 2014; Mourant et al., 2018; Vergu, Busson and Ezanno, 2010; Yashima and Sasaki, 2014; Tsai, Huang, Wen, Sun and Yen, 2011; Zhou, Xu, Hu, Yue, Li and Xia, 2020; Wardle et al., 2023; Lee et al., 2021; Long et al., 2018; Cao et al., 2022), and are highlighted here for the purpose of discussion. Some definitions were modified slightly from their original form to allow for direct comparisons between the models. Force of Infection options are highlighted in the table as they are the focus of this paper.

#### 2.1.1. Synthetic Data Generation

To verify geographical mixing dynamics separately from infection dynamics we examine different spatial population-distribution symmetries. We generated specific synthetic populations and examine the model outputs from deterministic infection processes as functions of grid-scale, and population spatial symmetry. Synthetic populations on square grids were generated to reflect spatial population variations that plausibly arise at the scale of 1 km.

#### 2.1.2. Spatial Interaction and Mixing Model

Regardless of model, disease present in cell *i* can cause infection in cell *j* with with a characteristic distance *s*_*d*_ during a defined time step (for example, 1 day). The “relative mixing” from cell *i* to cell *j* in each time step is specified using a discretized interaction kernel *κ*(*r*_*ij*_). This should be interpreted as the relative mobility, contact, or transmission between cell *i* and cell *j*, as is done in Mills and Riley (2014), Zhou et al. (2020), and Mourant et al. (2018). Figure 1 provides a visual example of how a diffusion kernel is discretized for the grid.

**Figure 1:**
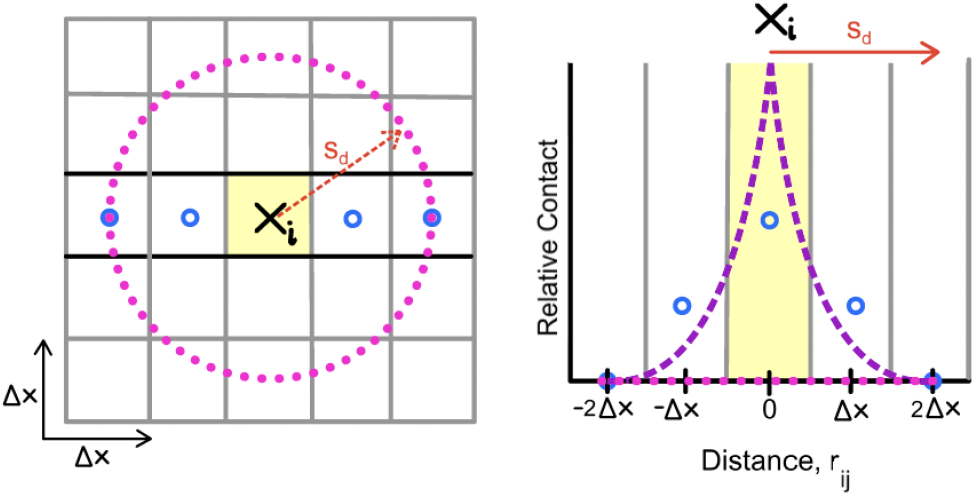
Left: A geographic grid with 25 cells. Δ*x* is the width of each cell. The characteristic radius of interaction is *s*_*d*_. Right: Illustration of the contact or probability of interaction *κ*(*r*_*ij*_) for some center-to-center distance, *r*_*ij*_, between cells *i* and *j*. The decay of the interaction kernel is shown in purple. The blue circles are the discretized geographical interaction, *κ*(*r*_*ij*_), between cell *j* and cell *i*.

Each entry *m*_*ij*_ of our mixing matrix *M* represents the average probability of mixing/interaction between the population from cell *i* with population in cell *j* (Wu et al., 2007; Mills and Riley, 2014; Zhou et al., 2020).

To ensure that *M* satisfies the definition of a mixing matrix (every row sums to 1), the entries *m*_*ij*_ must be normalized. Two methods are commonly used in the literature:

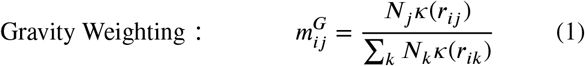

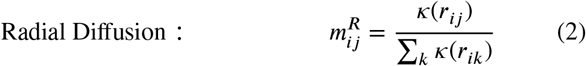

where *r*_*ij*_ is the distance between cell *i* and cell *j*. The first normalization method, 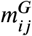, is the most common, which implies *gravity weighting*, where mixing is dependent on the underlying population densities *N*_*j*_. The second, 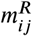, is simple *radial di*ff*usion*, where mixing is assumed to be *independent* of the relative population densities.

##### Remark 2.1

The factor 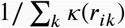 in Eqn (1) normalizes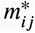, ensuring row sums add to 1. That is, the probability of contact between cell *i* and all other cells *j* including itself sums to 1, such that 100% of all possible contacts out of cell *i* are accounted for.

The interaction kernel *κ*(*r*_*ij*_) driving the mixing matrix *M* is the following:

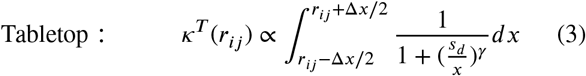

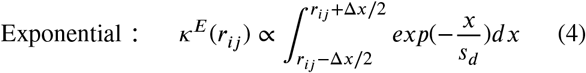

*κ*^*T*^ is the kernel used in Mills and Riley (2014) which resembles a tabletop assuming equal probability of contact below *s*_*d*_, while *κ*^*E*^ is the kernel used by Mourant et al. (2018), which is an exponential decay kernel similar to what is abstracted in Figure 1. In both cases, we assume *κ*(*r*_*ij*_) is the average value of the diffusion kernel in cell *j* relative to cell *i*, as visualized by the blue circle points in Figure 1. Parameter definitions for the mixing model are in Table 1.

In Mourant et al. (2018), *κ*(*r*) is interpreted as *transmission* between cells, and in Mills and Riley (2014), the same concept is used to model *movement* between cells, giving a similar picture of geographical dynamics in both cases. In neither case are long-term changes in population due to migration and birth included in the earlier work, nor do we consider it here.

For *κ*^*T*^, the more negative the mixing parameter *γ* is, the less the contact there is between cells. We used a fixed value of *γ* = −4 based off of the values proposed for use by Mills and Riley (2014).

#### 2.1.3. Infection Model

We construct a general SIR model on a grid, which we refer to as the *General Model* going forward. Then we further allow for two variations of this model based on common modeling decisions in the literature (Mills and Riley, 2014; Jones and Kulldorff, 2012; Mourant et al., 2018; Wu et al., 2007; Wardle et al., 2023; Pujante-Otalora et al., 2023; Lee et al., 2021; Long et al., 2018; Cao et al., 2022). All the possible modeling variations on the General Model explored here are summarized in Table 2.

Generally, cells of a population grid are identified by their location relative to the grid and the distribution of the population between infectious states. The following states are tracked within each cell: Susceptibles *S*_*i,t*_, Infecteds *I*_*i,t*_, Recovereds *R*_*i,t*_, and Total Population *N*_*i*_. Only Total Population remains constant throughout time, such that *N*_*i*_ = *S*_*i,t*_ + *I*_*i,t*_ + *R*_*i,t*_.

The general model is dependent on two main dynamics: infection within cells and between cells. The overall system of governing equations is then the following:

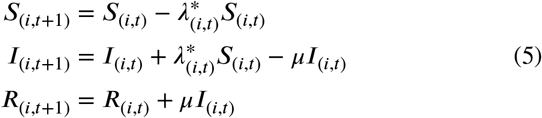

where 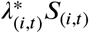 and *μI*_(*i,t*)_ represent all newly infected or recovered individuals in cell *i* at time *t* respectively.

The different *force of infection* 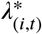 options commonly used in the literature are outlined in more detail in Section 3 and Table 2. Specifically, the verification and validation analysis performed here focuses on two different 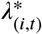, the Averaged Infection rate and the Per Capita Infection rate. See Equation (6) and Equation (7) for their respective definitions. Death is not modelled here nor is it part of the verification analysis because death reduces the total population. Similarly, birth and migration are not considered in this model; it is assumed that the timescale for disease progression is short enough to assume birth is negligible and the overall population remains relatively constant. Recovery could be added to this analysis though the usual dynamics in Eqn. 5 above.

In addition to these definitions for the force of infection, we will refer to a *well-mixed model*. The well-mixed infection model is Equation (5) such that 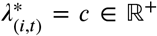 is constant, inferring infection is independent of spatial considerations.

### 2.2. Evaluation Metrics

We review the population data metrics and infection data metrics used to evaluate our results.

#### Basic Reproduction Number

Epidemiologically, the *basic reproduction number* ℛ_0_ is defined as the average number of secondary cases generated in a completely susceptible population in the limit of a very small number of initially infected cases at time *t* = 0 (van den Driessche and Watmough, 2008).

Mathematically, ℛ_0_ is a bifurcation parameter controlling the stability of the disease-free equilibrium. In the well-mixed SIR model, it is related to the non-parametric shape of the epidemic curve including the peak (when the change in infected cases first becomes zero, *İ* = 0) and final size of the epidemic (van den Driessche and Watmough, 2008). For a well-mixed SIR model, we can find: ℛ_0_ = *β*/*μ*.

#### Epidemic Peak

The *epidemic peak magnitude* is when the maximum of *I*, the infected population, is reached for some time *t*_*max*_, the *epidemic peak time*. We can calculate the time of the epidemic peak in a deterministic system with homogeneous population by finding *t*_*max*_ given *İ* = 0, *Ï* < 0 and subsequently *I*(*t*_*max*_). Numerically we can simply solve for max_*t*≥0_*I* and find the associated *t*_*max*_.

##### Remark 2.2

Our quantities of interest (ℛ_0_, epidemic peak(s), and epidemic peak time(s)) are often used in the literature and by public health professionals to characterize outbreaks. Therefore these will be used to compare different model outputs. For verification purposes, the epidemic peak magnitude and timings should be consistent across all geographic resolutions under certain conditions.

To verify the model, we check for a consistent final epidemic size (and therefore epidemic peak magnitude and timing) across all spatial resolutions and changing characteristic *s*_*d*_ for fixed *β* and *μ*. This process is explained in further detail in Section 3.1.1.

## 3. Results

We explored how the different modeling choices perform under a range of illustrative examples that cover: homogeneous populations, heterogeneous populations, and sparse populations.

To unravel how the geographic resolution of the population can bias the inferences and outbreak predictions we draw from a given model, we begin by evaluating how the modeling choices and parameters influence the epidemic curve, assuming we know the true parameter values. That is, if the underlying parameters are known (in some sense a best case scenario), how does the model prediction change as the geographic resolution of the population changes? as the underlying contact rates of the population change?

### 3.1. Impact of Infection Rate Decision

First we begin with the broad model choice decision of which infection rate formula to use. To understand how these infection rates differ, we show an explicit comparison of a 2-population grid.

#### 3.1.1. Infection Rate Assumptions

We will (a) first perform a comparison of the abstract modeling assumptions that generated the different force of infection rates, then (b) proceed with a mathematical verification process to check for mesh independence, to be defined.

##### Abstract Assumptions Verification

This verification process provides a direct comparison of guiding assumptions for the two geographically-resolved force of infection rates, 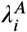 and 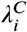, as defined in Table 2.

##### Model Consistency Verification (Mesh Independence Checks)

The spatiotemporal model results should converge to the single cell, well-mixed population result if the characteristic mixing distance *s*_*d*_ is large enough compared with the mesh size. That is, across all geographic resolutions of a given population, above a certain *s*_*d*_ threshold (to be determined), the cumulative epidemic curve and therefore final epidemic size should be consistent. In physics and engineering, this concept is referred to as *mesh independence*, where the mesh is what we refer to as the underlying population grid.

###### Remark 3.1

*Consistency* in spatiotemporal modeling guarantees that model results remain accurate and reproducible even when one or more input parameters are changed. We will explore consistency as it relates to changes in the underlying mobility and interactions of the population by fluctuating *s*_*d*_, or changing grid resolution. That is, model predictions should converge to the well-mixed model solution as interactions are increased or the grid resolution reduced, to prove that the model is consistent with the well-mixed model.

We explore consistency in the context of changes to the underlying interaction assumptions of the grid. That is, does the model continue to provide consistent results even as the underlying grid resolution or mixing assumptions change? This is especially important to consider as transmission is highly dependent on contact rates between individuals. Often, we want to model the expected infection trajectory *under the in*fl*uence of changing interaction dynamics*. To accurately model these changing interactions, we first must guarantee a consistent model result in the absence of such changes.

Mills and Riley (2014) verify that the model under the Averaged Infection rate 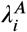 satisfies this consistency requirement in their Supplementary Text 1.

In the following **2-column *parallel* format**, we will (a) revisit the Mills and Riley (2014) proofs for Averaged Infection rate 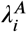 at left, and (b) rework their procedure for the Per Capita Infection rate 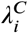 at right. This will allow us to confirm whether or not models under the Per Capita Infection rate assumption are consistent as well. **The text is deliberately spaced such that analagous portions of the proof appear roughly side by side**.

###### Remark 3.2

The proofs presented here use notation consistent with this paper, and have been further modified from what was originally presented by Mills and Riley (2014). In particular, the proof presented for Proposition 3.2A is a different approach than what was presented originally by Mills and Riley (2014).

#### 3.1.2. Abstract Assumptions Verification

##### Averaged Infection Rate 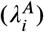

The force of infection or the average rate that individuals in cell *i* become infected, 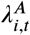, is the average of these actions per time step, given by (Mills and Riley, 2014; Wu et al., 2007; Wardle et al., 2023; Zhou et al., 2020):

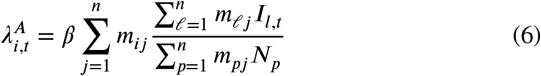

where *n* is the total number of cells in the grid, and for any cell *j, N*_*p*_ is the total number of individuals in cell *p, I*_*l,t*_ is the number of infected individuals in cell *l* at time step *t*, and infectious contacts are made with other individuals at a constant transmission rate *β*.

**According** to Mills and Riley (2014), this infection rate at which susceptible individuals in cell *i* become infected is dependent on the following: (1) their risk of infection from those infected in cell *i* already; (2) the risk of infection to susceptible individuals in cell *i* who traveled to *j*; and (3) the risk of infection from infected individuals in cell *j* who traveled to *i*.

**Comparing** the above equation to the defined paths of infection assumed to be handled, we can breakdown Equation (6) as the following:

- *βm*_*ii*_ is the total number of infectious contacts made by cell *i* with itself
- *βm*_*ij,i*≠*j*_ is the total number of infectious contacts made from cell *i* to other cells *j*
- 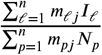 is the *average* fraction of contacts with infected individuals within cell *j* during the time step; this dimensionless value scales the relative infectivity of cell *j* to cell *i*; within this term:
  - *m*_ℓ*j*_ *I*_ℓ_ is the total number of *infectious* contacts made within the cell *j* (given *j* = ℓ), or from cell *l* traveling into cell *j* (given *j* ≠ ℓ); this represents all *infectious* contact from cells outside of cell *j* possibly into cell *j*;
  - *m*_*pj*_ *N*_*p*_ is the total number of contacts made within the cell *j* (given *j* = *p*), or from cell *p* traveling into cell *j* (given *j* ≠ *p*); this represents *all* contacts from cells outside of cell *j* possibly into cell *j* that cell *i* indirectly makes contact with.

**Thus**, we can verify that 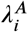 meets the model assumptions. □

##### Per-Capita Infection Rate 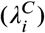

Alternatively 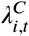 is the average of these actions per time step given by:

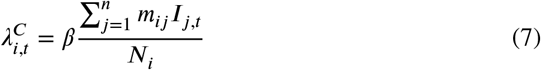

where *n* is the total number of cells in the grid, and for any cell *j, N*_*i*_ is the total number of individuals in cell *i, I*_*j,t*_ is the number of infected individuals in cell *j* at time step *t*, and infectious contacts are made with other individuals at a constant transmission rate *β*.

This generalization for the infection rate is more common in the literature for infection models that incorporate contact networks (Mourant et al., 2018; Long et al., 2018; Lee et al., 2021; Cao et al., 2022; Banos, Corson, Gaudou, Laperrière and Rey Coyrehourcq, 2015; Tsai et al., 2011; Vergu et al., 2010; Yashima and Sasaki, 2014).

**According** to Vergu et al. (2010), this rate represents “the probability an individual gets infected at time step *t* +1 given that they were susceptible at time step *t*.” Under 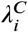, the force of infection generated from cell *i* contacting other cells *j* is distributed equally to the population of cell *i*, making it a *per capita* rate assumption. We will compare 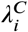 to the assumptions defined for 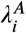 at left to emphasize the potential implications of this modeling choice.

**Comparing** this infection rate formulation to the dependency assumptions of 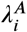, we find that 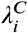 is dependent on the following:

- *βm*_*ii*_ is the total number of infectious contacts made by cell *i* with itself
- *βm*_*ij,i*≠*j*_ is the number of infectious contacts made from cell *i* to other cells *j*, not considering how cell *j* mixed with other cells

While the abstract assumptions built into this infection rate choice are strictly defined in the literature, by breaking down the formula it becomes clear that 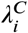fails to account for all possible types of mixing and infection that happen during a time-step in the same way that 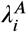 does. Specifically, infectious contact in cell *j* that happens during the same time step as infectious contact in cell *i* is not explicitly accounted for.

**Thus**, we can*not* verify that 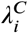 meets the model assumptions with the current information. □

#### 3.1.3. Model Consistency Verification

##### Proposition 3.1A

The basic reproduction number ℛ_0_ for SIR models under 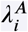 will be equivalent to the well-mixed SIR solution. That is, ℛ_0_ = *β*/*μ*.

*Proof*. Here we follow the proof originally done by Mills and Riley (2014) for the Averaged Infection rate 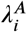.Note that the spectral radius ρ of the Next Generation matrix *G* for an SIR model is the basic reproduction number ℛ_0_, as defined in Section 2.2.

**We** seek to provide a simple expression for ℛ_0_ from the next generation matrix. Each cell of the Next Generation Matrix under 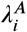 is defined as:

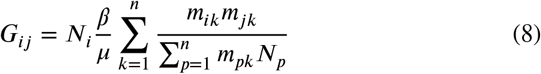

where *β* is the general rate of transmission per infectious contact, *μ* is the recovery rate, and *m*_*ij*_ is probability of interaction between cell *i* and cell *j* such that 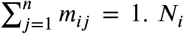 is the total number of individuals in cell *i*. Let 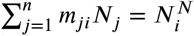, the total number of individuals in cell *i* during any time step. Then summing to find the *j*^*th*^ column of *G*, we find:

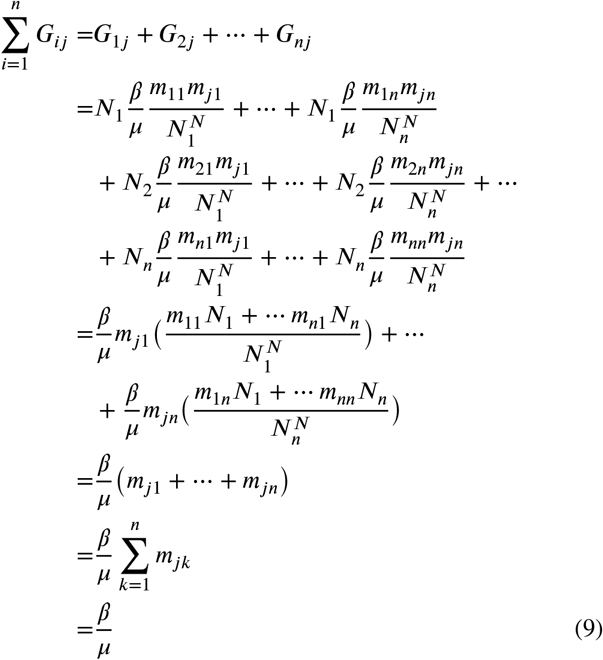

**This** holds for all columns *j* of G. Therefore it follows that ρ(*G*) = *β*/*μ*. This holds for any resolution and mixing matrix *m*_*ij*_.

Therefore, both 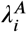 and the well-mixed SIR model will return the same ℛ_0_ estimate if both models have the same *β* and *μ* values. □

##### Proposition 3.1B

The basic reproduction number ℛ_0_ for SIR models under 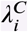 will be equivalent to models under the well-mixed model if the interaction matrix is symmetric. That is, ℛ_0_ = *β*/*μ* only if *m*_*ij*_ = *m*_*ji*_ ∀ *i, j* or *κ*(*r*_*ij*_) = *κ*_*j*_ ∀*i*.

*Proof*. Here we provide an abbreviated proof of how the same assumptions hold for the Per Capita Infection rate 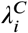.

**We** seek to provide a simple expression for ℛ_0_ from the next generation matrix. Each cell of the Next Generation Matrix under 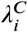 is defined as:

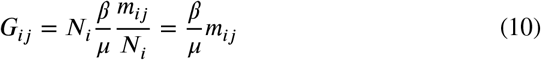

where *β* is the general rate of transmission per infectious contact, *μ* is the recovery rate, and *m*_*ij*_ is probability of interaction between cell *i* and cell *j* such that 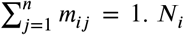 is the total number of individuals in cell *i*. Let 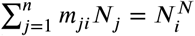, the total number of individuals in cell *i* during any time step. Then summing to find the *j*^*th*^ column of *G*, we find:

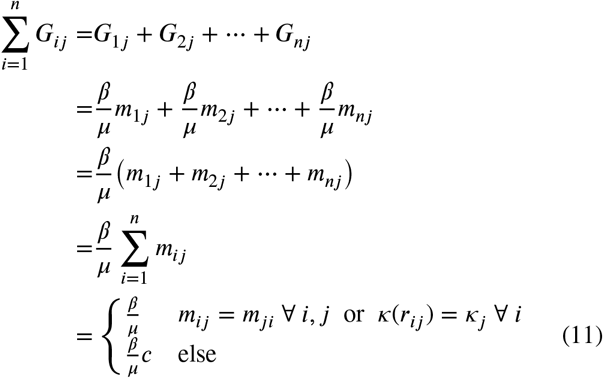

where *c* ∈ ℝ^+^ s.t. generically *c* ≠ 1. Note if *m*_*ij*_ = *m*_*ji*_ ∀ *i, j*, we can use the row sums conclusion 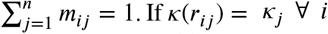 we can guarantee that the column sums add to 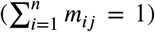 as shown in Proposition A.1 and A.2 of the Appendix.

**This** holds for all columns *j* of G. Therefore it follows that ρ(*G*) = *β*/*μ*. This holds for any resolution and mixing matrix *m*_*ij*_, if *m*_*ij*_ = *m*_*ji*_ ∀ *i, j* or *κ*(*r*_*ij*_) = *κ*_*j*_ ∀ *i*.

##### Proposition 3.2A

Therefore, both 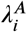 and the well-mixed SIR model will return the same ℛ_0_ estimate if both models have the same *β* and *μ* values, and *m*_*ij*_ = *m*_*ji*_ ∀ *i, j* or *κ*(*r*_*ij*_) = *κ*_*j*_ ∀ *i*.

##### Proposition:3.2A

The final epidemic size will be consistent for any arbitrary grid resolution or mixing condition under *λ*^*A*^ as long as *m*_*ij*_ > *ϵ* ∀*i, j*, given some *ϵ* << 1. That is, the final epidemic size for SIR models under *λ*^*A*^ will be the same as the final epidemic size for well-mixed SIR models.

***Proof***. Our metapopulation SIR model is as defined in Equation (5) where *μ* is the recovery rate, and now we explore 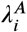 as the force of infection in cell *i*:

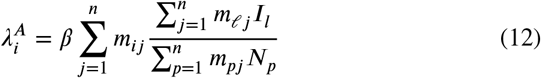

given *n* is the total number of cells on the grid and *m*_*ij*_ is our mixing model from cell *i* to cell *j*. Again, *N*_*i*_ is the total number of individuals in cell *i*. Transmission occurs during a interaction event between an infected and a susceptible individual at some rate *β*.

**Because** there is no way to replenish susceptibles, a final equilibrium must exist, which we will label *S*_*i*_(∞), *I*_*i*_(∞), and *R*_*i*_(∞), such that *I*_*i*_(∞) = 0 and *N*_*i*_ = *S*_*i*_(∞) + *R*_*i*_(∞). Further, we have already shown that ℛ_0_ = *β*/*μ* for this system.

In each cell *i*, the fraction who did not get infected will be denoted σ_*i*_ = *S*_*i*_(∞)/*S*_*i*_(0), such that the final epidemic size in each cell will be *x*_*i*_ = *N*_*i*_(1 − σ_*i*_). Then the total final epidemic fraction of the entire population is:

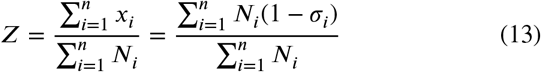

**We** divide 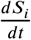 from Equation (5) by *S*_i_ and integrate from 0 to, then we have:

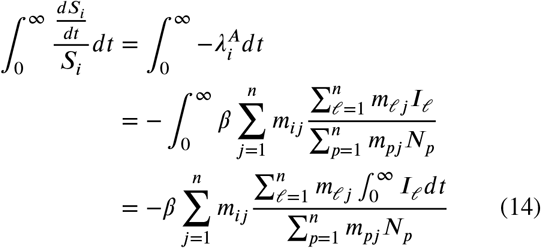

and note that:

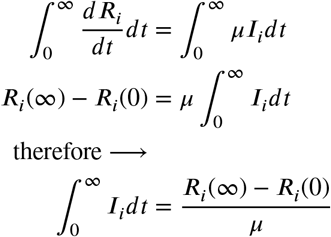

##### Proposition 3.2B

The final epidemic size generically will *not* be consistent for any arbitrary grid resolution or mixing condition under *λ*^*c*^ as long as *m*_*ij*_ > *ϵ* ∀*i, j* given some *ϵ* << 1, unless the population is spatially homogeneous. Unless the population is homogeneous, the final epidemic size for SIR models under *λ*^*c*^ will *not* be the same as the final epidemic size for well-mixed SIR models.

***Proof***. Our metapopulation SIR model is as defined in Equation (5) where *μ* is the recovery rate, and now we explore 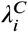 as the force of infection in cell *i*:

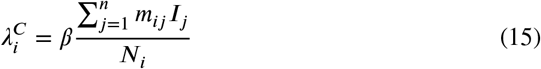

given *n* is the total number of cells on the grid and *m*_*ij*_ is our mixing model from cell *i* to cell *j*. Again, *N*_*i*_ is the total number of individuals in cell *i*. Transmission occurs during a interaction event between an infected and a susceptible individual at some rate *β*.

**Because** there is no way to replenish susceptibles, a final equilibrium must exist, which we will label *S*_*i*_(∞), *I*_*i*_(∞), and *R*_*i*_(∞), such that *I*_*i*_(∞) = 0 and *N*_*i*_ = *S*_*i*_(∞) + *R*_*i*_(∞). Further, we have already shown that ℛ_0_ = *β*/*μ* for this system, under certain conditions.

In each cell *i*, the fraction who did not get infected will be denoted σ_*i*_ = *S*_*i*_(∞)/*S*_*i*_(0), such that the final epidemic size in each cell will be *x*_*i*_ = *N*_*i*_(1 − σ_*i*_). Then the total final epidemic fraction of the entire population is:

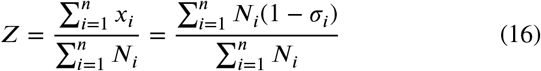

**We** divide 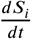 from Equation (5) by *S*_i_ and integrate from 0 to∞, then we have:

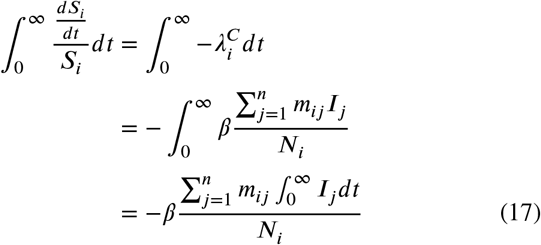

and note that:

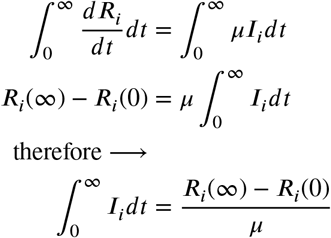

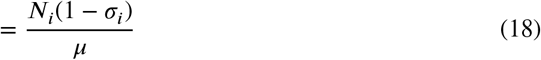

since *R*_*i*_(∞) − *R*_*i*_(0) = *x*_*i*_ = *N*_*i*_(1 − σ_*i*_), the final epidemic size in cell *i*. Therefore:

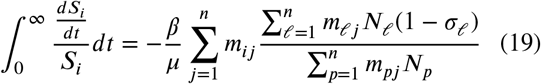

**Recall** from Ma and Earn (2006) that it is also true that:

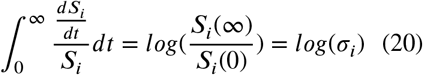

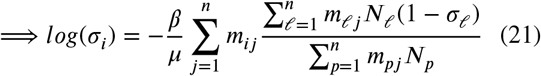

And equivalently since *x*_*i*_ = *N*_*i*_(1 − σ_*i*_):

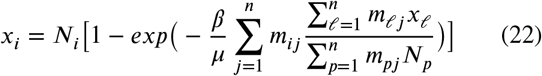

With the condition that *m*_*ij*_ > *ϵ* ∀ *i, j* for some *ϵ* << 1, then if the final epidemic size is non-zero, *x*_*i*_ or (1 − σ_*i*_) is also non-zero. To demonstrate this, consider an arbitrary cell *i* with no infection such that σ_*i*_ = 1.

This would imply:

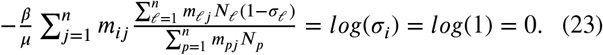

**Imposing** *m*_*ij*_ > *ϵ* ∀ *i, j* for some *ϵ* << 1, for the LHS to be zero then (1 − σ_*i*_) = 0∀*i*, implying no cell has an infection. So by our assumptions on *m*_*ij*_, if *Z* > 0 then *x*_*i*_ > 0 for all cells *i*.

Then it follows:

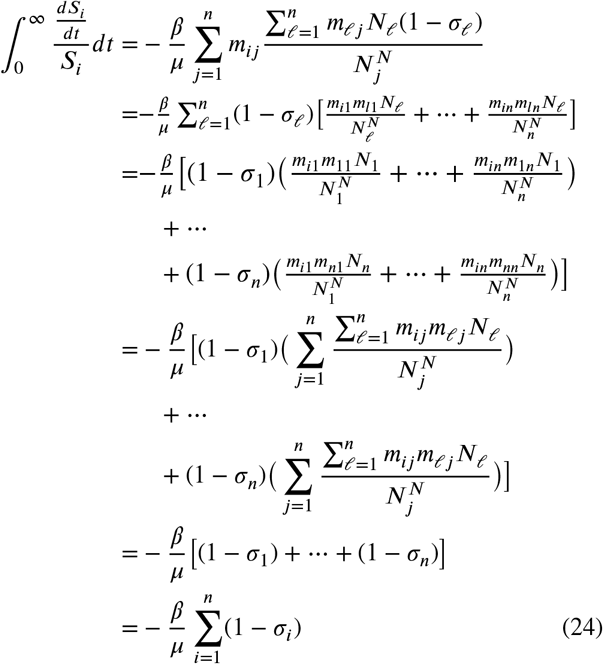

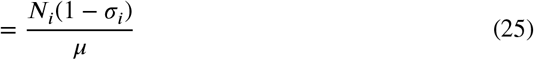

since *R*_*i*_(∞) − *R*_*i*_(0) = *x*_*i*_ = *N*_*i*_(1 − σ_*i*_), the final epidemic size in cell *i*. Therefore:

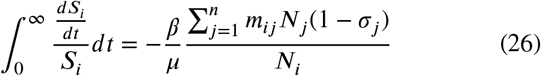

**Recall** from Ma and Earn (2006) that it is also true that:

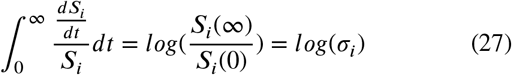

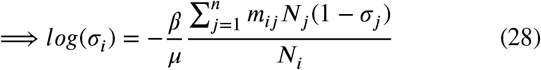

And equivalently since *x*_*i*_ = *N*_*i*_(1 − σ_*i*_):

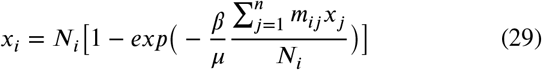

With the condition that *m*_*ij*_ > *ϵ* ∀ *i, j* for some *ϵ* << 1, then if the final epidemic size is non-zero, *x*_*i*_ or (1 − σ_*i*_) is also non-zero. To demonstrate this, consider an arbitrary cell *i* with no infection such that σ_*i*_ = 1.

This would imply:

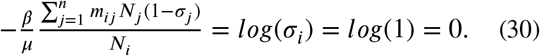

**Imposing** *m*_*ij*_ > *ϵ* ∀ *i, j* for some *ϵ* << 1, for the LHS to be zero then (1 − σ_*i*_) = 0∀*i*, implying no cell has an infection. So by our assumptions on *m*_*ij*_, if *Z* > 0 then *x*_*i*_ > 0 for all cells *i*.

Then it follows:

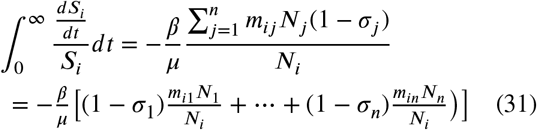

No further simplification for Equation (31) is possible without further information about the spatial structure of the underlying population; that is we need more information about *m*_*ij*_ and *N*_*i*_ ∀ *i, j*. Consider two possibilities:

a. the population is homogeneous and *N*_*i*_ = *N*_*j*_ ∀ *i, j*
b. the population is heterogeneous and ∃ *i, j* such that *N*_*i*_ ≠ *N*_*j*_

We can rewrite Equation (31) if *m*_*ij*_ > *ϵ* ∀ *i, j*:

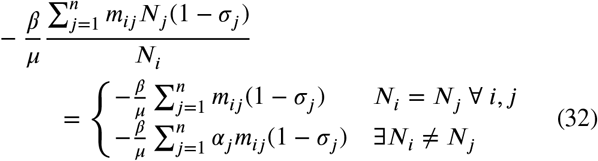

where *α*_*j*_ = *N*_*j*_/*N*_*i*_ ∈ ℝ^+^.

**Then** from Equation (22) and (24), we can show that the final epidemic size is given implicitly by:

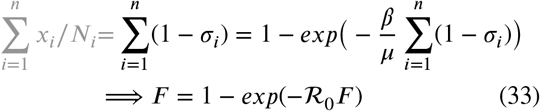

where *F* is the vector of final epidemic sizes for each cell *i*. This is consistent with the calculations for the well-mixed SIR model (Ma and Earn, 2006). Further, note that this final formula does not depend on any of the spatially dependent elements from our original equation.

**The** final epidemic size will be consistent across different characteristic distance values, given that *m*_*ij*_ > *ϵ* for some *ϵ* << 1. □

**Then** from Equation (29) and (32), we can show that the final epidemic size is given implicitly by:

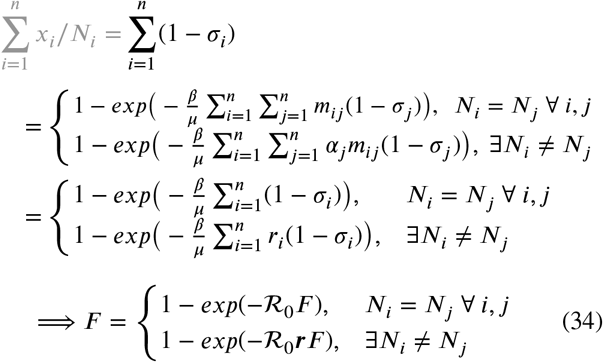

where 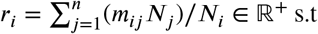 s.t. generically *r*_*i*_ ≠ 1 for all *m*_*ij*_ and *N*_*j*_, and *F* is the vector of final epidemic sizes for each cell *i*.

We can see for the homogeneous case where *N*_*i*_ = *N*_*j*_∀ *i, j*, we recover the final epidemic size equation as seen in Ma and Earn (2006) for the well-mixed SIR model, and for 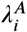 (shown at left). Heterogeneous cases where ∃*N*_*i*_ ≠ *N*_*j*_ generate an extra term ***r*** that is generically not equal to 1, and therefore we can*not* recover the original final epidemic size equation.

**The** final epidemic size will generically *not* be consistent across different characteristic distance values, even beyond the threshold alluded to in Mills and Riley (2014) if the population is not homogeneous. □

#### 3.1.4. Comparing the Proofs

##### Remark 3.3

Mills and Riley (2014) origianlly use *m*_*ij*_ > 0 to ensure contact between all cells *i* and *j*. We found that this condition was insufficient to guarantee consistency during numerical tests, and instead chose to impose the stricter condition of *m*_*ij*_ > *ϵ* ∀ *i, j* given some *ϵ* << 1.

In conclusion, the Per Capita Infection rate 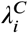 generically cannot return constant epidemic size predictions for *all* populations, grid resolutions and characteristic distance *s*_*d*_, under the realistic assumption of a non-heterogeneous population. Further, the Per Capita Infection rate cannot even guarantee a consistent basic reproduction unless the mixing matrix satisfies *m*_*ij*_ = *m*_*ji*_ ∀ *i, j*. This is problematic as ℛ_0_ is a common metric used by public health professionals for characterizing an epidemic outbreak and determining potential risk to the population of interest.

In Appendix A.3 we give several numerical examples illustrating the potential simulation results when *r* ≠ 1 if the population is not homogeneous. Further, in Figure 3, Figure 8, and Figure 9 we can see that under 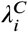, increasing *s*_*d*_ greater than some threshold will continually decrease the expected final epidemic size. That is, we can visually verify that the model under 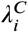 will not converge to the well-mixed model solution even as its parameters tend towards a well-mixed model.

Both infection rate assumptions are consistent for homogeneous populations, in which case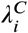 provides a computationally cheaper option. For heterogeneous populations, 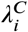 is *not* guaranteed to return consistent results. Equivalently we can say that this model is never mesh-independent. Therefore, models made under the Per Capita Infection rate 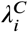 are not verifiable with regards to consistent epidemic size predictions. Models built under the Averaged Infection rate 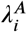 however are verifiable and spatially consistent.

### 3.2. Impact of Mixing Model on Infection Dynamics

Since the lowest resolution (single cell grid) is equivalent to assuming uniform mixing of the population, we can infer that high *s*_*d*_ values should mimic this assumption. All of our consistency checks are to verify that under *well-mixed* conditions (e.g. *m*_*ij*_ > *ϵ, ϵ* << 1) the model converges to the solution under the well-mixed SIR model.

Figure 3-9 demonstrate a visual argument of consistency to support the proofs presented in Section 3.1.2 and 3.1.3. The following can be seen in these figures:

- *Left column:* this shows the population grid for a given spatial resolution, with the cell that was initially infected (marked by an “x”).
- *Center column:* this shows for **fixed spatial resolution** as seen at left, the fi*nal epidemic size* in blue versus *different interaction assumptions* (in this case, increasing the characteristic distance *s*_*d*_ parameter which is used to define the mixing matrix *m*_*ij*_). Vertical lines represent proposed thresholds on *m*_*ij*_, specifically note in grey where *m*_*ij*_ > 0 (threshold recommended by Mills and Riley (2014)) versus where *m*_*ij*_ > *ϵ* (our recommended threshold). Further, 3 thresholds in shades of pink are defined for comparison in the far right column. With this column, we can compare if the model provides consistent final epidemic size predictions for different underlying interaction assumptions.
- *Right column:* this shows for **fixed interaction assumptions** the fi*nal epidemic size* versus *different spatial resolutions*. The colors of each plot in this column correspond to the colored vertical lines in the center column. Now we can compare if the model provides consistent final epidemic size predictions for different spatial resolutions.

#### 3.2.1. Basic Reproduction Number (ℛ0)

The basic reproduction number ℛ_0_ is a commoanly used metric by public health professionals to determine the potential severity of an outbreak. Model estimates for ℛ_0_ under infection rate 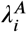 will be consistent across spatial resolutions and different mixing models.

However, model estimates for ℛ_0_ under infection rate 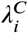 will *not* be consistent across spatial resolutions and different mixing models, unless specific conditions are met. Namely, that either the mixing matrix is symmetric (*m*_*ij*_ = *m*_*ji*_ ∀ *i, j*), or that the underlying interaction assumption between cells and their neighbors are symmetric for all cells (*κ*(*r*_*ij*_) = *κ*_*j*_ ∀ *i*). This is a highly restrictive assumption. For interaction rates of rural versus urban communities, this assumption may not hold.

#### 3.2.2. Resolution of the Grid (Δx)

When the spatial resolution of the population grid is lowered (or equivalently Δ*x* is increased), the heterogeneous population density patterns become homogeneous. Under 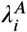,this has no effect on the epidemic size or epidemic peak magnitude. The changing resolution does change the predicted epidemic peak timing.

Under 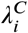, changing the grid resolution changes the epidemic size, peak magnitude, and peak timing predicted by the model, even when all other parameters are held constant. This is consistent with the results for the ℛ_0_ proof, Proposition 3.1B, as ℛ_0_ is related to the final epidemic size and therefore peak timing and magnitude.

#### 3.2.3. Symmetry of the Mixing Matrix

Radial normalization of the mixing matrix 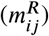 was selected not only because it is used in the literature, but also because it generates a symmetric mixing matrix. Gravity weighting normalization of the mixing matrix 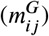 generates an asymmetric mixing matrix that favors more mixing in more densely populated cells.

As can be seen in the right column of Figure 2, symmetric mixing matrices are more sensitive to the choice of infection rate 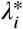.Per Capita infection rate 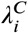, which does not assume mixing is weighted by relative populations, diffuses and becomes negligible as *s*_*d*_ increases under the symmetric mixing assumption. Therefore, while ℛ_0_ may be consistent under symmetric mixing assumptions for 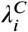 (as found with Proposition 3.1B), the *epidemic peak timing* and *magnitude* are not guaranteed to be consistent for any population. All the epidemic curves in Figure 2(d) should have the same ℛ_0_ value, but present with very different predictions for the realized epidemic curve.

**Figure 2:**
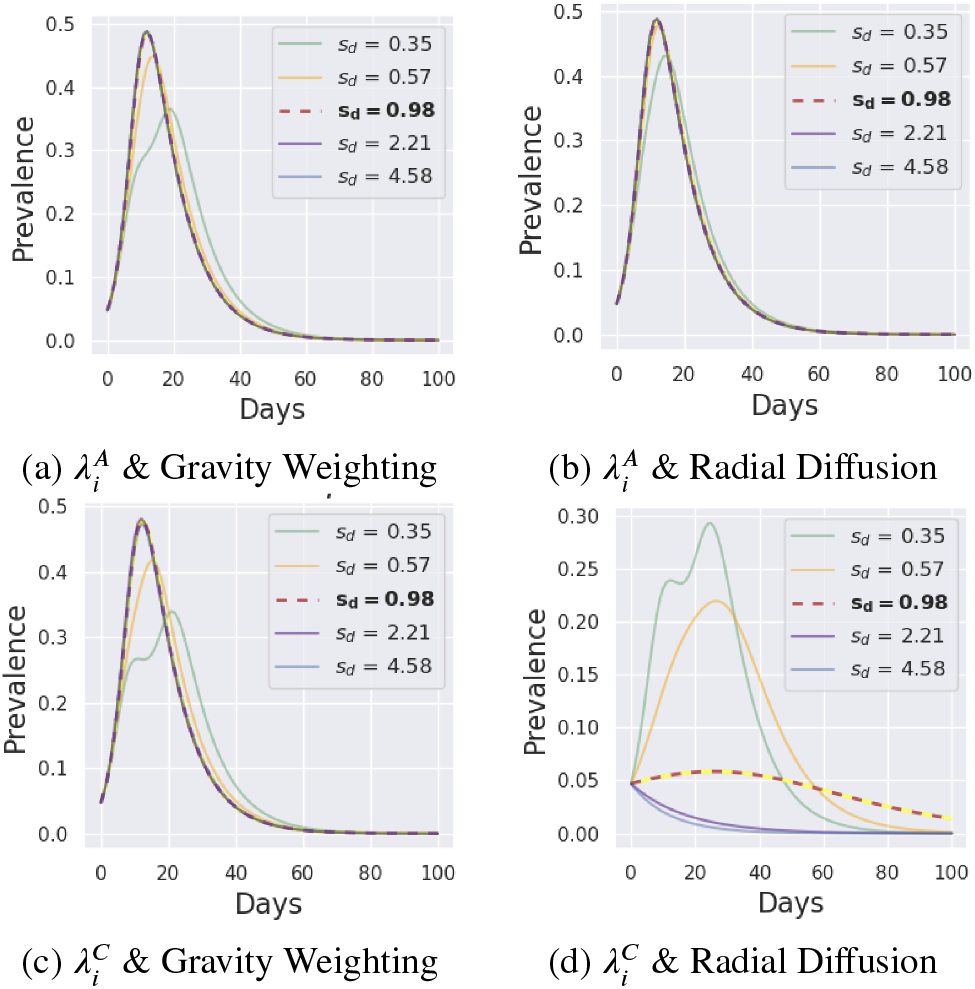
Comparison of sample infection curves for *κ*^*T*^ (Tabletop kernel) and 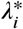 on a sparse population. This simulation used *β* = 0.5 and *μ* = 0.1 on a sparse 2-cell population shown in Figure 13 (Bottom).

As we learned in Proposition 3.2B, models built under 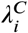 will only be guaranteed consistent final epidemic sizes if the population is homogeneous. This effect is not obvious in Figure 2(c), but is extremely clear under a symmetric mixing matrix as seen in Figure 2(d). In this way, symmetric mixing matrices can be used as a first pass consistency test for models.

#### 3.2.4. Characteristic Distance sd

##### Averaged Infection Rate 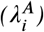

Across all resolutions and *β* values, reducing *s*_*d*_ delayed epidemic peak timing and reduced epidemic peak magnitude for 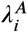.This effect becomes more prominent as the resolution on the grid increases. This general trend is emphasized in Figure 3(a), and can be seen for other general population grid selections in Figure 7.

**Figure 3:**
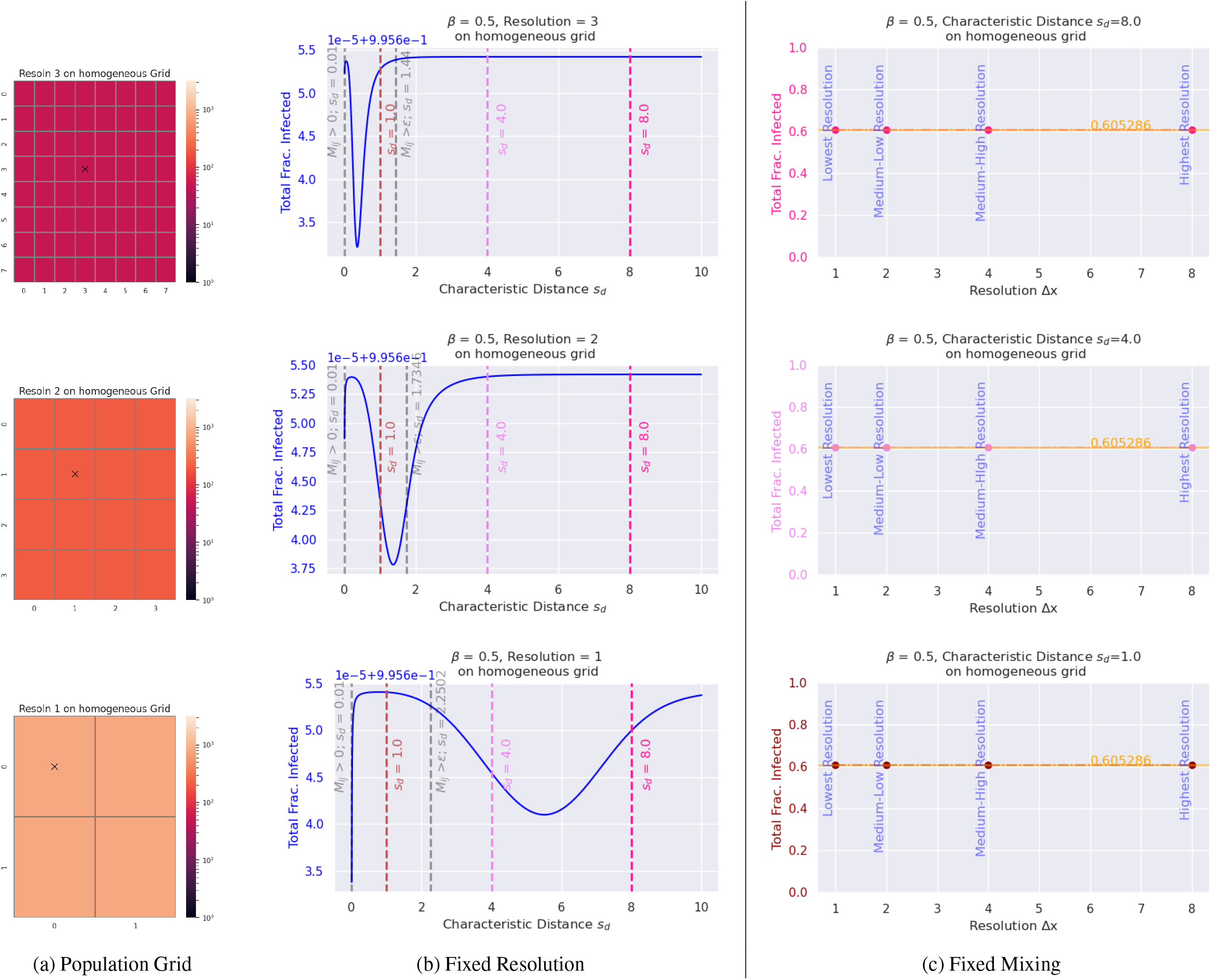
Comparison of consistency with respect to the final epidemic size (aka total fraction infected) when the resolution (b) or the characteristic distance *s*_*d*_ (c) are fixed respectively. The yellow line is the expected final epidemic size from the well-mixed model. Vertical red/pink lines in the middle column (b) correspond to the fixed *s*_*d*_ values highlighted in the far right column (c). Results shown here are for **Averaged Infection rate** 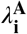, **radial diffusion normalization** of *κ*^*T*^, and *β* = 0.5, *μ* = 0.1.

The characteristic distance *s*_*d*_ has no impact the epidemic peak timing nor magnitude for the lowest resolution (single cell) populations. Since no neighboring cells exist to interact with, the contact model and therefore *s*_*d*_ have no impact on the infection dynamics.

##### Per Capita Infection Rate 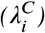

For 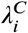, decreasing *s*_*d*_ can *increase* epidemic peak magnitude and decrease epidemic peak timing for sparse populations under radial normalization of mixing, as emphasized in Figure 5(b) and the right hand plot of Figure 2(b). All other mixing model combinations under 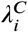 yield the same results as was observed for 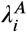.

**Figure 4:**
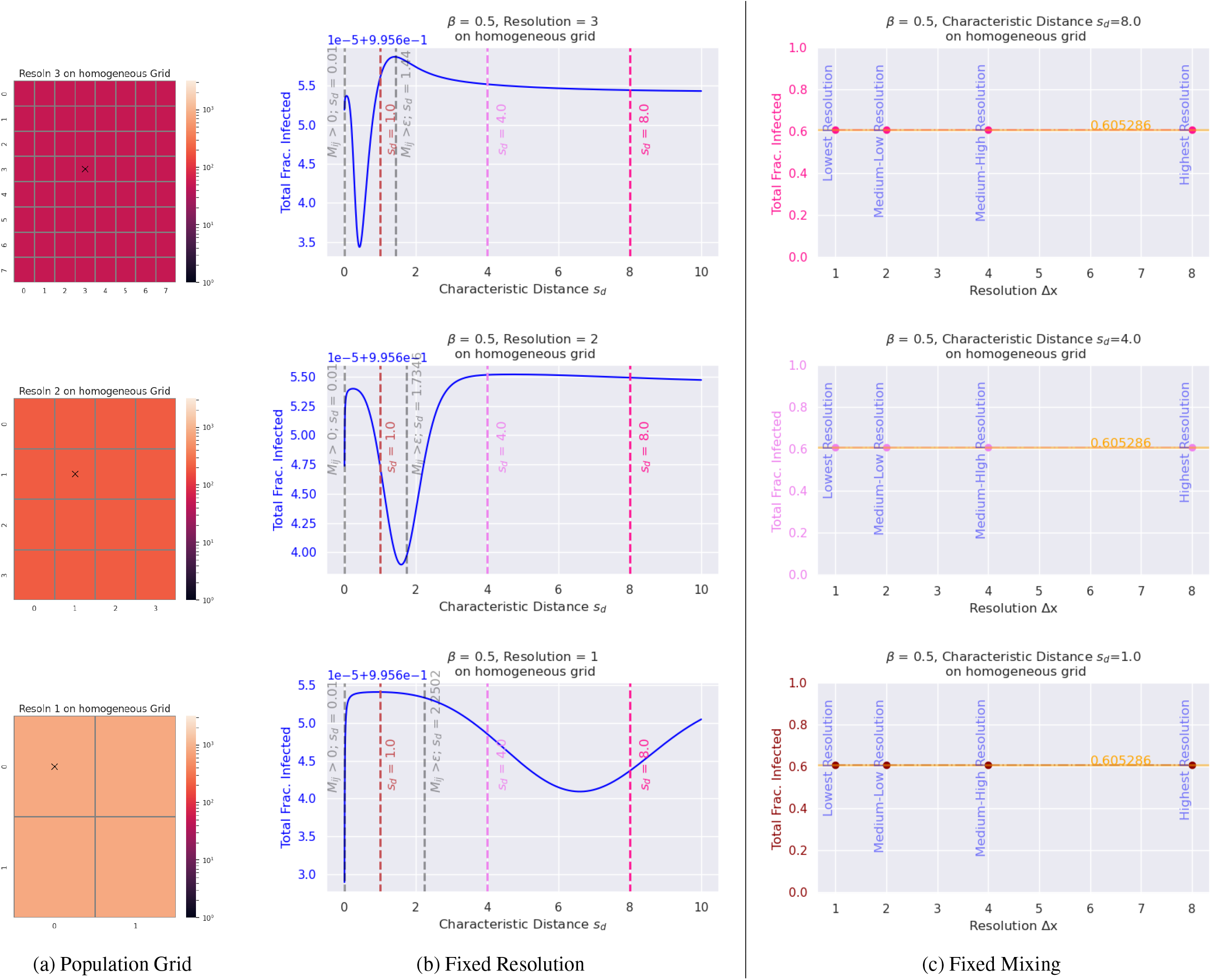
Comparison of consistency with respect to the final epidemic size (aka total fraction infected) when the resolution (b) or the characteristic distance *s*_*d*_ (c) are fixed respectively. The yellow line is the expected final *s*_*d*_ values highlighted in the far right column (c). Results shown here are for **Per Capita Infection rate** 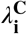, **radial diffusion normalization** of *κ*^*T*^, and *β* = 0.5, *μ* = 0.1.

**Figure 5:**
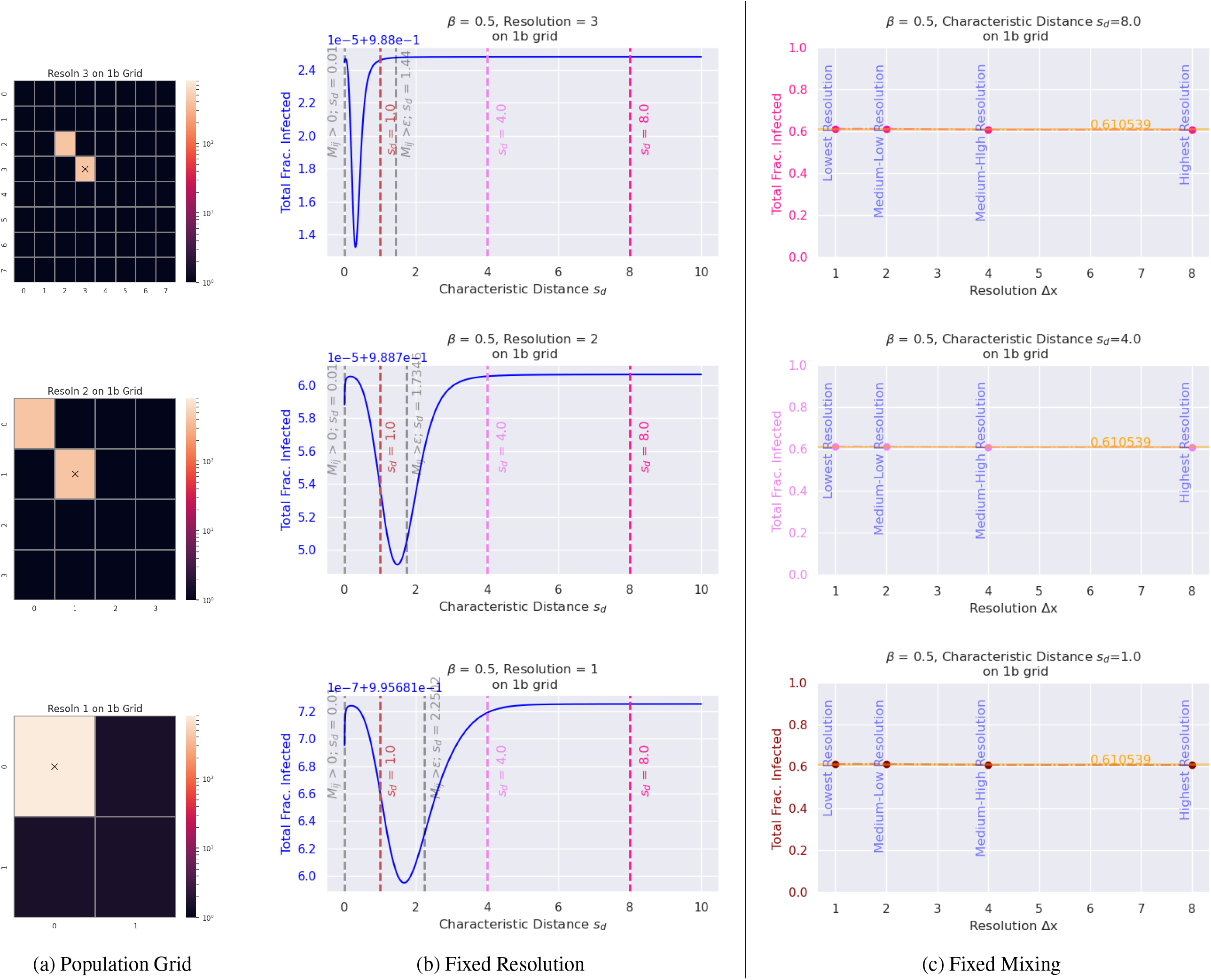
Comparison of consistency with respect to the final epidemic size (aka total fraction infected) when the resolution (b) or the characteristic distance *s*_*d*_ (c) are fixed respectively. The yellow line is the expected final epidemic size from the well-mixed model. Vertical red/pink lines in the middle column (b) correspond to the fixed *s*_*d*_ values highlighted in the far right column (c). Results shown here are for **Averaged Infection rate** 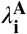, **radial diffusion normalization** of *κ*^*T*^, and *β* = 0.5, *μ* = 0.1.

## 4. Discussion

In the Section 3, we defined verifiable model assumptions, consistency conditions, and the potential implications these have on modeling efforts. Here we clarify and breakdown based on different modeling goals which model choices are appropriate when, as well as suggested guidelines for future “best practice”.

### 4.1. Generating Consistent Epidemic Size

#### Averaged Infection Rate 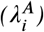

Mills and Riley (2014) proved mathematically in their Supplementary Text S1 that under the Averaged Infection rate 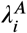,the final epidemic size (the final cumulative attack rate) and the basic reproduction number for a population was the same at all resolutions and all characteristic distances above a certain threshold.

This result holds during our verification process, as we can see in Figure 3, Figure 5, and Figure 7 that if *m*_*ij*_ > *ϵ* ∀ *i, j*, the total fraction infected (and therefore epidemic size) appears to be consistent across all grid resolutions. Note in all of the aforementioned figures, in grey a threshold for *m*_*ij*_ > 0 ∀ *i, j* and *m*_*ij*_ > *ϵ* ∀ *i, j* are provided for comparison. *m*_*ij*_ > 0 ∀ *i, j* was the original threshold suggested by Mills and Riley (2014), but as we can see visually, is insufficient to guarantee consistency for any resolution and mixing condition. We found *ϵ* = 0.001 to be generally sufficient.

#### Per Capita Infection Rate 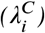

For the Per Capita Infection rate 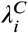, not all conditions generate consistent epidemic peak sizes, as we found in Section 3.1.1. In Figure 4, the constant epidemic size assumption holds, as it is a homogeneous population. But, for heterogeneous populations as seen in Figure 6, Figure 8 and Figure 9, constant epidemic size assumption clearly fails to hold. The final epidemic size for a population is *not* the same across all characteristic distances and resolutions, no matter the threshold.

**Figure 6:**
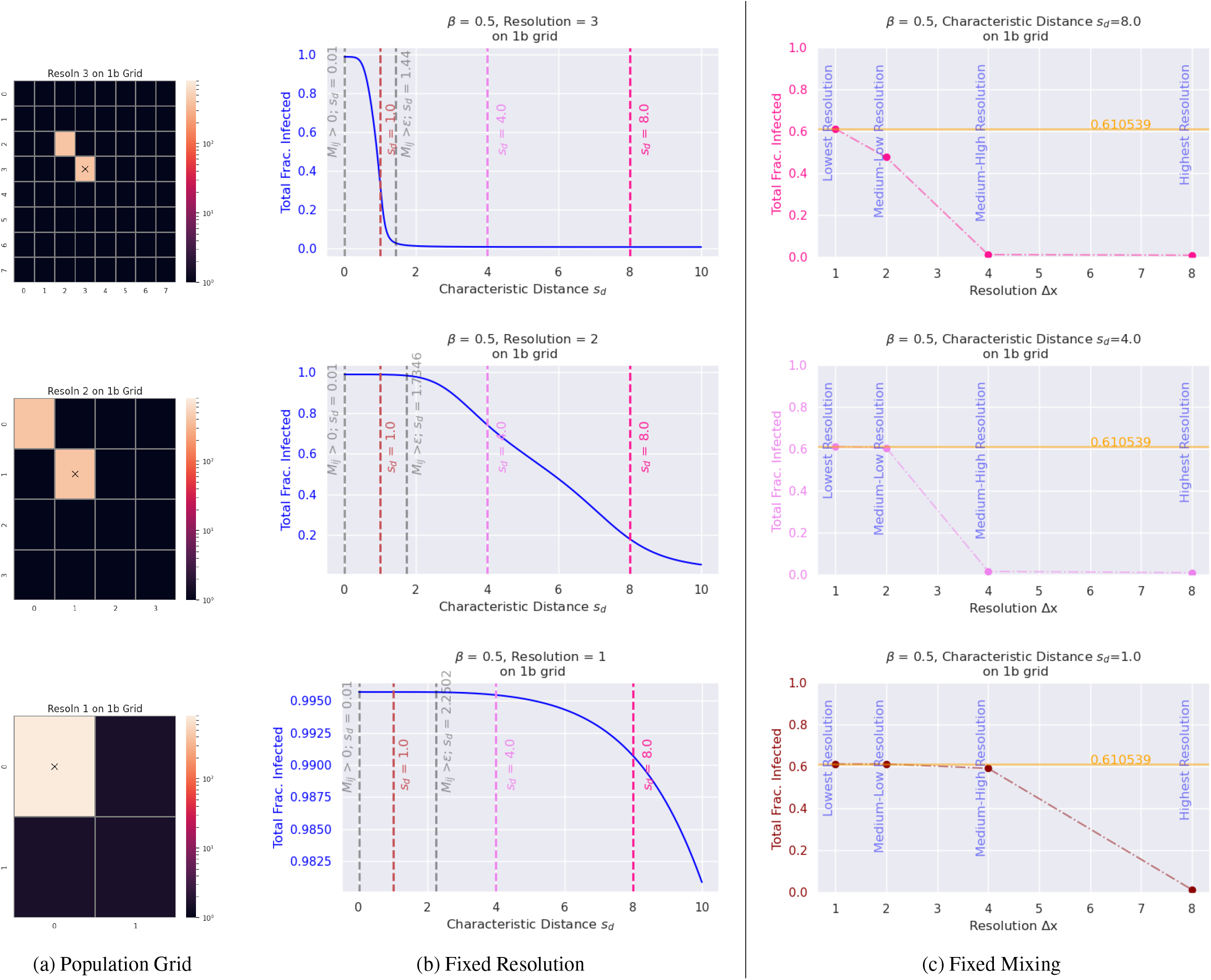
Comparison of consistency with respect to the final epidemic size (aka total fraction infected) when the resolution (b) or the characteristic distance *s*_*d*_ (c) are fixed respectively. The yellow line is the expected final epidemic size from the well-mixed model. Vertical red/pink lines in the middle column (b) correspond to the fixed *s*_*d*_ values highlighted in the far right column (c). Results shown here are for **Per Capita Infection rate** 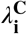, **radial diffusion normalization** of *κ*^*T*^, and *β* = 0.5, *μ* = 0.1.

**Figure 7:**
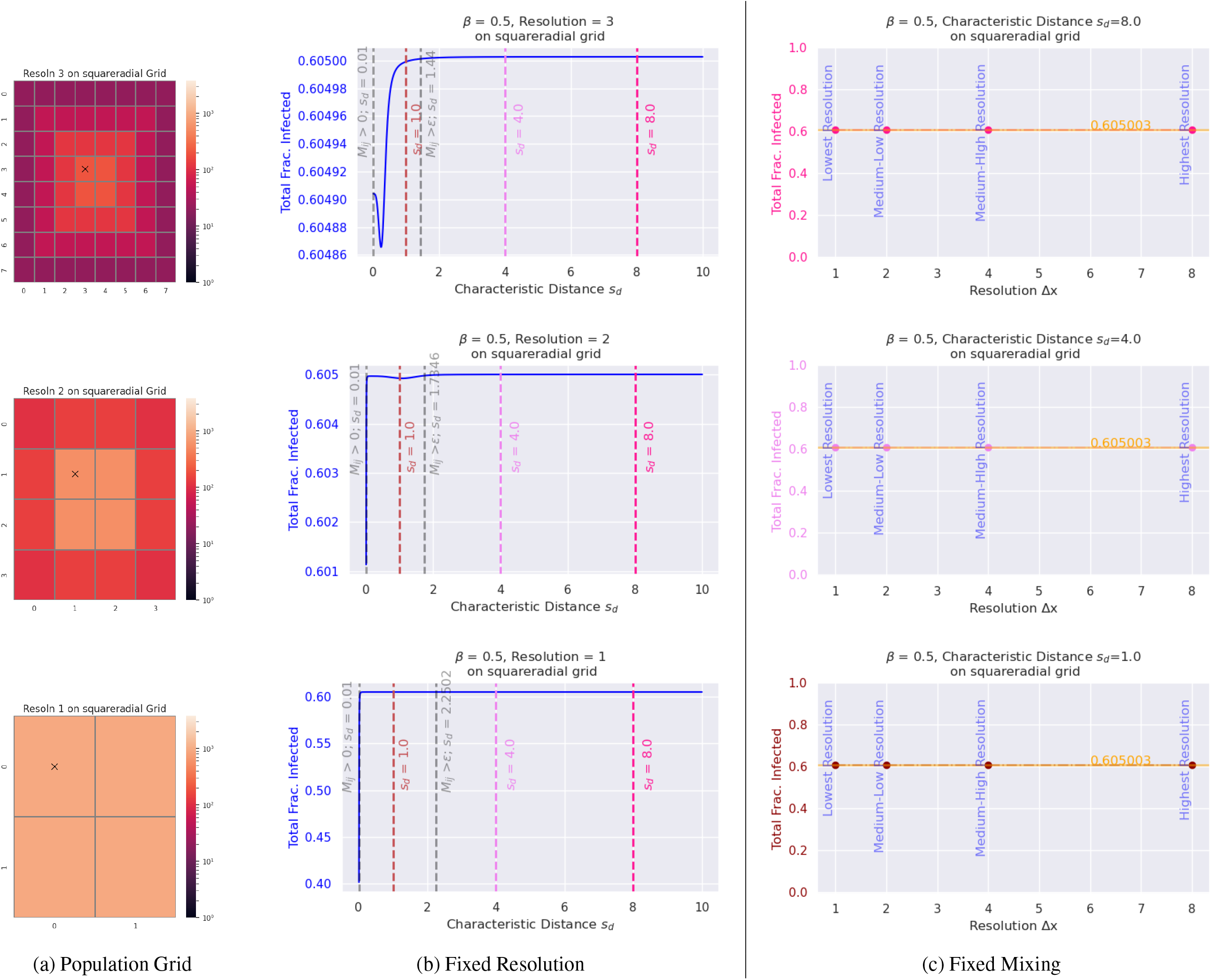
Comparison of consistency with respect to the final epidemic size (aka total fraction infected) when the resolution (b) or the characteristic distance *s*_*d*_ (c) are fixed respectively. The yellow line is the expected final epidemic size from the well-mixed model. Vertical red/pink lines in the middle column (b) correspond to the fixed *s*_*d*_ values highlighted in the far right column (c). Results shown here are for **Averaged Infection rate** 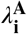, **radial diffusion normalization** of *κ*^*T*^, and *β* = 0.5, *μ* = 0.1.

**Figure 8:**
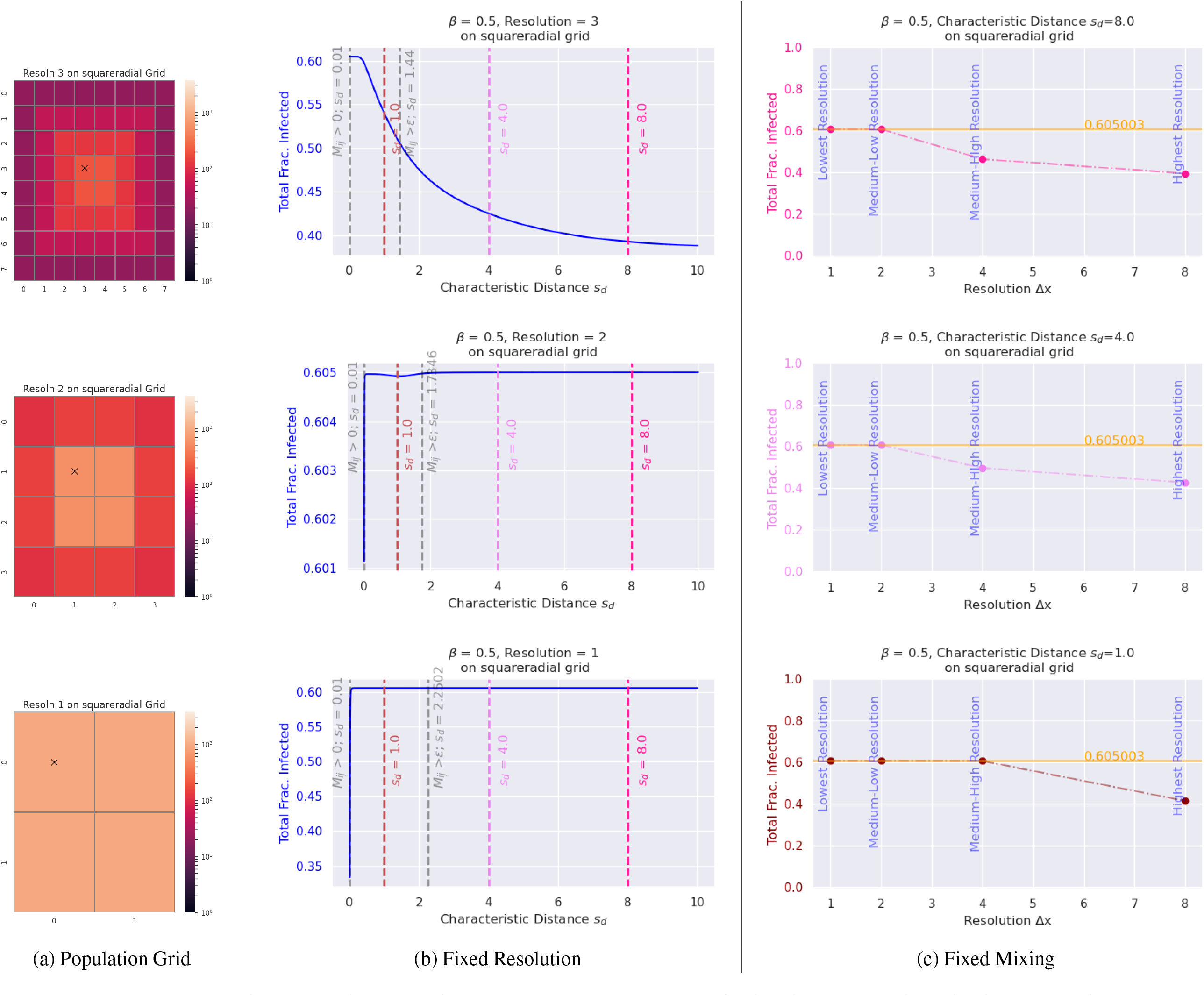
Comparison of consistency with respect to the final epidemic size (aka total fraction infected) when the resolution (b) or the characteristic distance *s*_*d*_ (c) are fixed respectively. The yellow line is the expected final epidemic size from the well-mixed model. Vertical red/pink lines in the middle column (b) correspond to the fixed *s*_*d*_ values highlighted in the far right column (c). Results shown here are for **Per Capita Infection rate**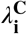, **radial diffusion normalization** of *κ*^*T*^, and *β* = 0.5, *μ* = 0.1.

**Figure 9:**
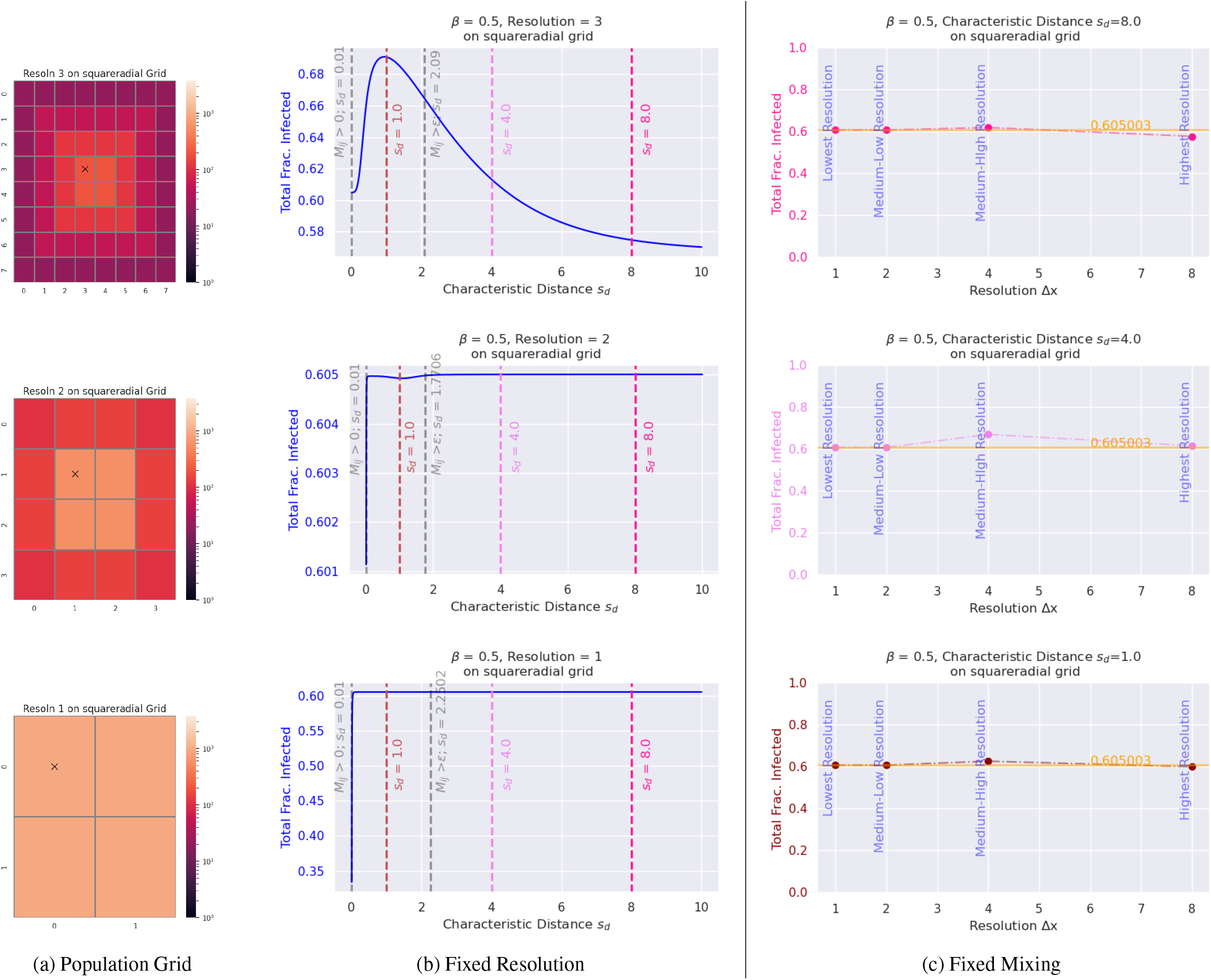
Comparison of consistency with respect to the final epidemic size (aka total fraction infected) when the resolution (b) or the characteristic distance *s*_*d*_ (c) are fixed respectively. The yellow line is the expected final epidemic size from the well-mixed model. Vertical red/pink lines in the middle column (b) correspond to the fixed *s*_*d*_ values highlighted in the far right column (c). Results shown here are for **Per Capita Infection rate**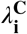, **gravity weighting normalization** of *κ*^*T*^, and *β* = 0.5, *μ* = 0.1.

Further, for Figure 6, Figure 8 and Figure 9, the model implications do not match what we expect from reality as increasing the mixing radius *s*_*d*_ results in a decreasing total fraction infected and therefore smaller final epidemic size.

We noted under 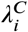 and radial normalization, as *s*_*d*_ *decreases* the total fraction infected of the epidemic peak increased. This is due to a ‘leaking’ effect on the infection rate if the relative population of cells is not accounted for properly. The relative infection rate of cells diffuses more rapidly on the grid as *s*_*d*_ increases, thereby decreasing the realized infection rate in the populated cell under radial normalization. This results in the prediction that for wide enough mixing kernels, the epidemic will fail to start, as seen for the blue and purple curves in Figure 2(d) and further emphasized in the summary consistency plots in Figure 6(b). Therefore, in effect *increasing* mixing on this grid under these assumptions *decreases* risk of infection.

Additionally, due to the *per capita* assumption implied by the 1/*N*_*i*_ term, 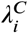 is highly sensitive to the particular population density of a cell or region that it describes. Lower populated cells are expected to express relatively higher infection rates than higher populated cells due to the *per capita* assumption. As such, 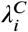 will be sensitive to changes in geographic resolution for heterogeneous populations.

Models built under the 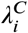 assumption are expected to give different predictions, nowcasts, and forecasts depending on the population grid resolution and mixing assumptions for heterogeneous or sparse populations. Consistent results are only guaranteed for homogeneous populations, such as those seen in Figure 3.

### 4.2. Finite Size Effects: Capturing Mitigation Efforts Accurately

Finite size effects are errors that arise when infinite systems are modeled with finite units. In this case, our consistency tests in Section 3.1.1 were to verify that under no mitigations and a well-mixed population, our macro-level dynamics perform as expected for characteristic distance *s*_*d*_ above a certain threshold. We recommend finding Δ*x* such that one can satisfy *m*_*ij*_ > *ϵ* ∀ *i, j*.

Below the threshold, new dynamics in the model results appear: lower epidemic peak magnitude, broader infection curves, and later epidemic peak timing. When the characteristic distance *s*_*d*_ < Δ*x*, the size of the cell, even stranger dynamics can occur: multiple epidemic peaks.

Researchers want to accurately model the impact of mitigation methods such as vaccination, new treatments, or mobility restrictions. These mitigations may be happening at different geographic scales, and to accurately capture their potential changes to the system, an adequate geographic resolution of the population needs to be selected.

At a minimum, the mitigation dynamics should be greater than the minimum cell size. That is, if a mitigation reduces the characteristic mixing distance *s*_*d*_ (0) to some smaller *s*_*d*_ (1), then both *s*_*d*_ (0), *s*_*d*_ (1) > Δ*x* to prevent finite size effects.

#### 4.2.1. Lockdown and Movement Restrictions

Lockdown procedures and canceled flights were common mitigations during the COVID-19 outbreak (Pearson, Colombo, Cecchini and Scarpetta, 2020; Lai, Ruktanonchai, Carioli, Ruktanonchai, Floyd, Prosper, Zhang, Du, Yang and Tatem, 2021), but movement restrictions during outbreaks have a much longer history: Roadblocks and airport checkpoints during the Ebola 2014 West Africa outbreak (Bausch and Rojek, 2016; Bardosh, Leach and Wilkinson, 2016), livestock movement restrictions during foot-and-mouth disease, rinderpest, or swine flu outbreaks (Tildesley, Brand, Brooks Pollock, Bradbury, Werkman and Keeling, 2019; Mourant et al., 2018; Dixon, Sun and Roberts, 2019; Ferdousi, Moon, Self and Scoglio, 2019), or temporarily closing live animal markets to reduce potential zoonotic infectious contacts (Yu, Wu, Cowling, Liao, Fang, Zhou, Wu, Zhou, Lau, Guo et al., 2014; Xiao, Newman, Buesching, Macdonald and Zhou, 2021).

All of the above examples include movement restrictions at different geographic scales, from global all the way down to specific locations. Researchers should first consider the minimum characteristic distance *s*_*d*_ they want to model. Where unrestricted movement may have a radius of 7km, this same movement under lockdown procedures may be predicted to reduce to 3km. Then a grid with *less than* Δ*x* = 3km should be selected for accurate modeling. Given standard data availability, in this case selecting a grid with Δ*x* = 1km would suffice.

##### Remark 4.1

Ensuring the model meets the cell-size threshold offers multiple benefits beyond just minimizing finite size effects in the output. Models that satisfy the cell-size criteria will produce consistent outcomes even when a finer geographic grid is used. This approach allows modelers to manage computational costs without compromising the robustness of the model. By ensuring the model is spatially consistent, modelers are better able to deliver stakeholders reliable, accurate, and reproducible results.

#### 4.2.1. Overestimating the Number of Epidemic Peaks

Mills and Riley (2014) mentioned that for *s*_*d*_ below an undefined threshold, the number of epidemic peaks could be overestimated by the model. As we noted above, this is an example of a finite size effect.

To prevent an overestimation of epidemic peaks in the epidemic model output and guarantee consistent epidemic size regardless of resolution or mixing assumptions, we recommend a general threshold of *s*_*d*_ > Δ*x*. This threshold is sufficient, as can be seen in any of Figure 3, Figure 5, or Figure 7.

Under the Per Capita Infection rate 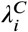, this same threshold will also guarantee that multiple peaks are not generated. Consistent epidemic size however is not possible under this assumption, as explained in Section 3.1.1.

##### Choosing the Model

On homogeneous population grids, when ∃*i, j* s.t. *m*_*ij*_ < *ϵ*, the Per Capita Infection rate 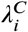 predicts infection spreading slower, and therefore reaching a lower epidemic peak later than what is predicted under the Averaged Infection rate 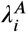 (See Figure 11 in the Appendix).

**Figure 10:**
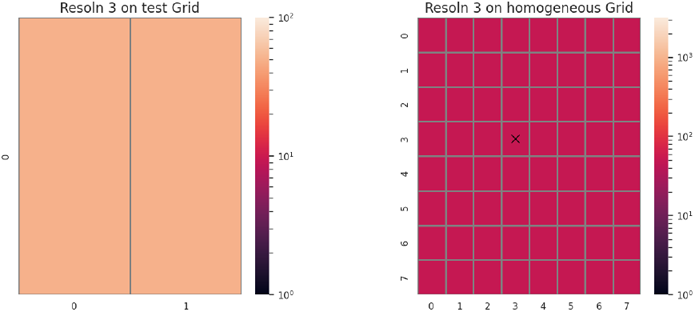
Homogeneous grid populations. Left: Grid with (37) two cells where Δ*x* = 1km. Right: 8×8 population grid.

**Figure 11:**
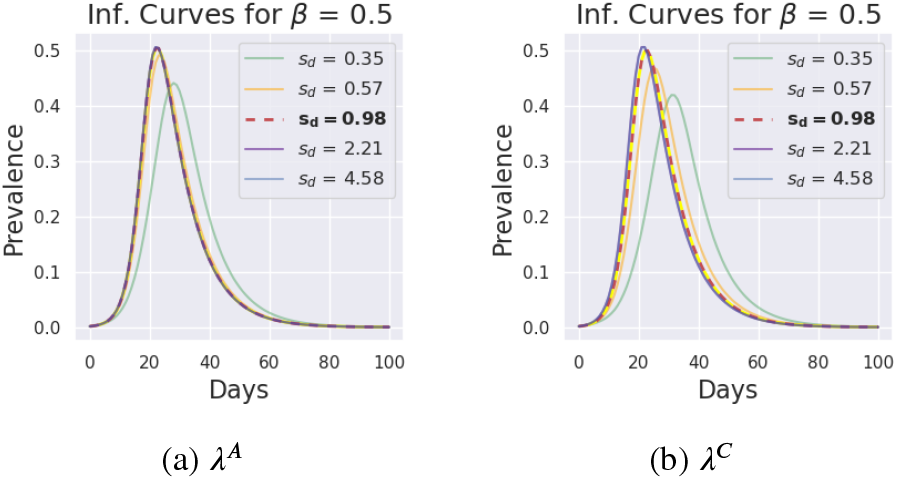
Comparison of infection curves for *κ*^*T*^ (Tabletop Kernel) and gravity weighting for a homogeneous population. This simulation used *β* = 0.5 and *μ* = 0.1, with varying characteristic distance *s*_*d*_. Note the epidemic peak is reached at the same time under 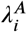 and 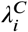 for all *s*_*d*_ > 1 choices are the same. For *s*_*d*_ < 1km, the modeling results begin to differ, with 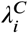 being more sensitive to small *s*_*d*_ values.

As was noted before, 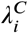 under radial normalization of the mixing matrix implies the reverse: decreasing *s*_*d*_ increases the total fraction infected. This implication that decreasing mixing increases risk of infection does not match what has been observed in reality. Predictions, nowcasts, and forecasts made from models under these assumptions should be carefully evaluated before being used for policy and resource allocation decisions.

Predictions generated under these conditions for 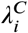 would potentially give policy makers a false sense of security that infection could be dramatically slowed at a faster rate than what 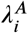 would imply.

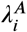 gives consistent general predictions that for any resolution, mixing normalization, and interaction kernel choice: decreasing *s*_*d*_ (reducing interaction radius) decreases the total fraction of the population infected and potentially increases the epidemic peak timing (see Figure 3(b), Figure 5(b), or Figure 7(b) for reference).

## 5. Conclusions

While the Per Capita Infection rate 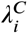 is the more common infection rate modeling choice in the literature compared to the Averaged Infection rate 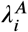,it does not directly account for differing population ratios. As such, this infection rate is highly sensitive to the normalization choice and the interaction assumptions of the mixing matrix, as well as heterogeneous populations. It cannot produce verifiable, consistent results for *every* population grid at *any* resolution, and performs especially poorly on sparse populations.

The *Per Capita* infection rate assumption fails the consistency tests for heterogeneous populations and non-symmetric mixing assumptions. The largest numerical errors occur in the limit of lowest symmetry, whether as sparse geographical distribution of the population or preferential travel to highly-populated locations. Future modeling efforts in spatiotemporal disease modeling should be wary of this limitation, particularly when working with more heterogenous or less dense populations.

In the post-COVID-19 pandemic modeling era, it is important that verifiable, validated models are presented to policy makers. Future spatiotemporal outbreak modeling work should demand models be consistent across resolutions and mixing choices at a minimum to guarantee reproducible, verifiable results.

## Data Availability

All data produced in the present work are contained in the manuscript

## 6. Acknowledgements

This work is the result of the research funded by the National Science Foundation (grant no. DMS-1853032), the Clare Boothe Luce Program for Women in STEM, the Michigan Institute for Computational Discovery and Engineering, Los Alamos National Laboratory Centers, and DTRA funding through CBCall18-CBI-07-2-0011.

This work was made possible in part by the cooperative agreement CDC-RFA-FT-23-0069 from the CDC’s Center for Forecasting and Outbreak Analytics. Its contents are solely the responsibility of the authors and do not necessarily represent the official views of the Centers for Disease Control and Prevention.

We thank Dr. Judith Mourant for first pointing out the symptoms of grid-dependent model results arising from per capita normalization. We also have special thanks for Dr. Sara del Valle for her assistance and advice during the early stages of this work.

## A. Appendix

### A.1. Row and Column Sums for Radial Diffusion

#### Proposition A.1A

The row sums for the radial diffusion normalization method of the mixing matrix 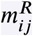 will always add to 1 given any arbitrary interaction kernel *κ*^∗^(*r*_*ij*_). However, the same cannot be generically guaranteed for the column sums of 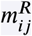.

*Proof*. Recall the general formula for the radial diffusion normalization is the following:

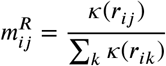

Then the sum of row *i* is the following:

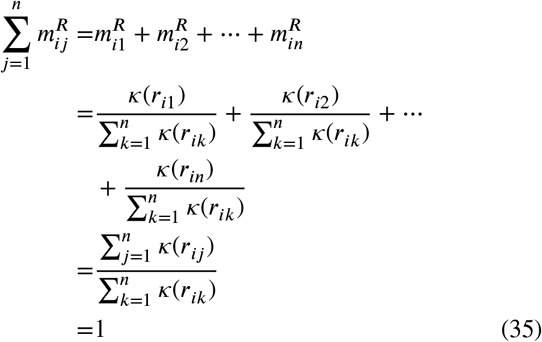

Therefore the row sums of 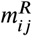 will always equal 1, regardless of the underlying interaction kernel.

The sum of column *j* is then the following:

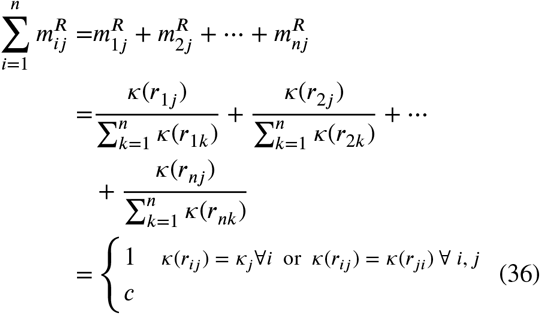

where *κ*_*j*_ ∈ ℝ^+^, and *c* ∈ ℝ^+^ s.t. generically *c* ≠ 1.

Unlike with the row sums of 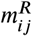,we cannot generically prove that the column sums will also always add to 1. A simple counter example can be considered using the following: let there be a two-cell system. Let *κ*(*r*_11_) = 0.3, *κ*_12_ = 0.7, *κ*_21_ = 0.4, and *κ*_22_ = 0.8. Then the mixing matrix under *m*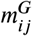 would be defined as the following:

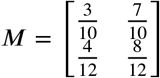

It is trivial to see in this counter example that while the row sums are 1, the column sums are not. Therefore, we cannot generically prove that the columns sums of mixing matrices generated under 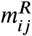 will add to 1.

### A.2. Row and Column Sums for Gravity Weighting

#### Proposition A.2A

The row sums for the gravity weighting normalization method of the mixing matrix 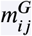 will always add to 1 given any arbitrary interaction kernel *κ*^∗^(*r*_*ij*_). However, the same cannot be generically guaranteed for the column sums of 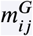.

*Proof*. Recall the general formula for the radial diffusion normalization is the following:

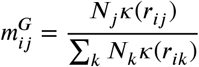

Then the sum of row *i* is the following:

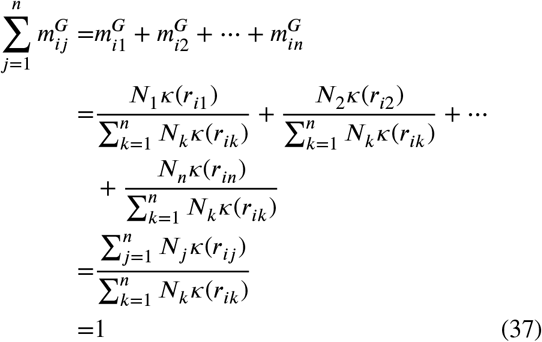

Therefore the row sums of 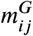 will always equal 1, regardless of the underlying interaction kernel.

The sum of column *j* is then the following:

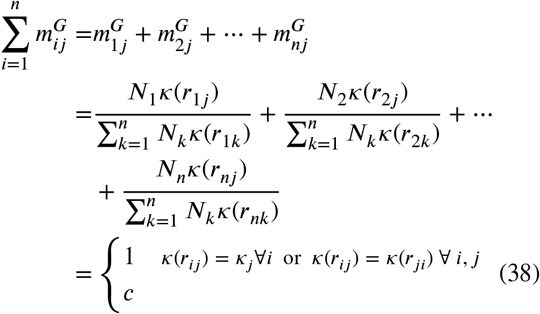

where *κ*_*j*_ ∈ ℝ^+^, and *c* ∈ ℝ^+^ s.t. generically *c* ≠ 1.

Unlike with the row sums of 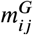, we cannot generically prove that the column sums will also always add to 1. A simple counter example can be considered using the following: let there be a two-cell system such that *N*_1_ = 50 and *N*_2_ = 200. Let *κ*(*r*_11_) = 0.3, *κ*_12_ = 0.7, *κ*_21_ = 0.4, and *κ*_22_ = 0.6. Then the mixing matrix under 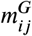 would be defined as the following:

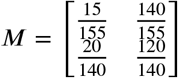

It is trivial to see in this counter example that while the row sums are 1, the column sums are not. Therefore, we cannot generically prove that the columns sums of mixing matrices generated under 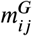 will add to 1.

### A.3. Examples

To better understand the potential impacts of modeling under the Per Capita Infection rate 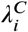 assumption on heterogeneous populations, we provide two explicit examples of a sparsely populated grid and a densely populated but heterogeneous grid.

As 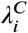 is a very common modeling assumption in the literature, we provide here explicit examples of where and how this assumption fails, specifically in comparison to the Averaged Infection rate 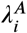, which we have shown above to be consistent across geographic resolutions and increasing characteristic mixing distances *s*_*d*_.

#### Homogeneous Populations

First, we start with a 2 cell example where the mathematical implications can be explicitly explored, and then confirm that for a large grid (in this case 8 by 8 cells) both 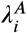 and 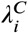 return consistent results as predicted.

##### 2 Equal Cells

Assume there are two populated cells, 0 and 1, and no other cells on the grid, as visualized in Figure 10. Under these conditions the mixing matrix normalization choice is not relevant, and therefore is not discussed.

Averaged Infection Rate Analysis: Under 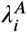 the *S*_(*i,t*+1)_ equation can be explicitly expressed as the following:

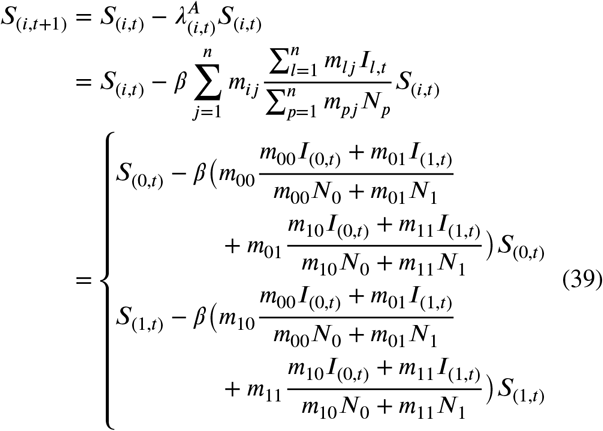

For large *N*_0_ = *N*_1_ and a single infection in cell 0 (that is, *I*_(0,0)_ = 1), *s*_*d*_ = 1km, these equations above become under 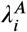:

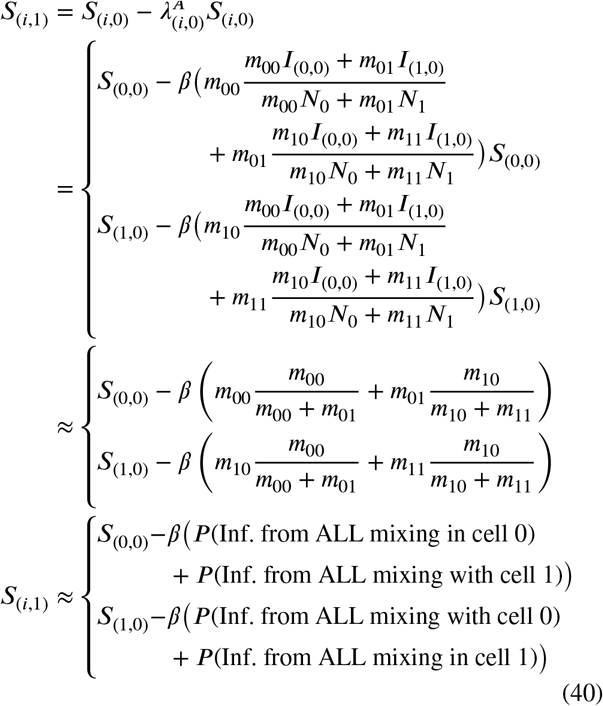

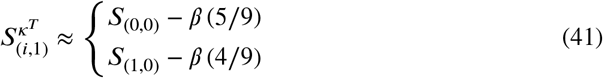

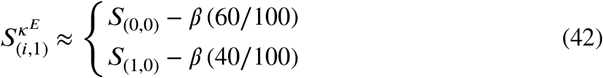

Equation (40) assumes that possible infectious interactions from cell *i* to *j* is relative to the total fraction of possibly infectious contacts in cell *j*. This accounts for all types of interactions/mixing happening simultaneously during the time step.

Using the Tabletop interaction kernel *κ*^*T*^, we can approximate the relative infection rates of cell 0 and cell 1 to be Equation (41) for the first time-step. Using the Exponential interaction kernel *κ*^*E*^, they instead approximate to be Equation (42).

Per Capita Infection Rate Analysis: Repeating this exercise for 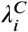, the *S*_(*i,t*+1)_ equation can be explicitly expressed as the following:

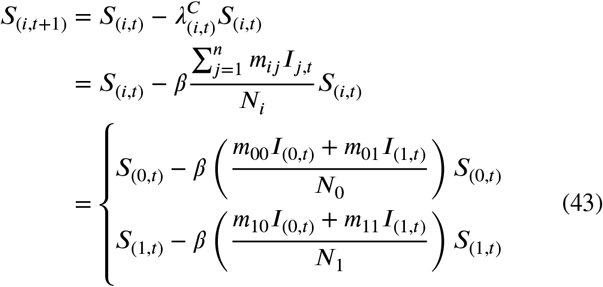

And for large *N*_0_ = *N*_1_ and a single infection in cell 1 (that is, *I*_(1,0)_ = 1), *s*_*d*_ = 1km, under 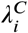:

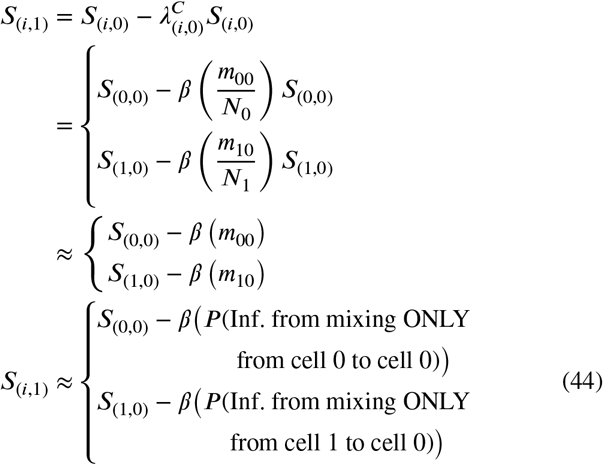

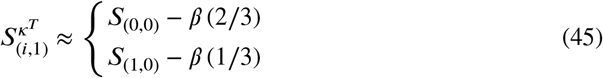

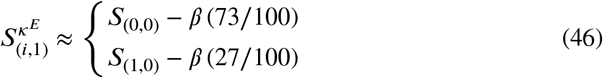

Equation (44) assumes that infection is determined only by interacting with the initially infected cell, cell 1. Other types of interactions/mixing are not accounted for under this formulation.

Examining the driving Equation (43), infection in cell 0 only considers mixing due from cell 0 to 1, and not infection that may have been introduced due to mixing from cell 1 into 0. Infection as a result spreads slower across a grid than it would under 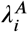,allowing more time for individuals to recover before they can reintroduce infection, and therefore resulting in a lower total fraction infected and later peak time for infection curves.

The direct effect of this choice is a more extreme ratio between the relative infection of cell 0 versus cell 1 than what was observed under 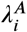,As before, Equation (45) approximates the relative infection rates of cell 0 and cell 1 under the Tabletop interaction kernel *κ*^*T*^, while Equation (46) does this under the Exponential interaction kernel *κ*^*E*^ for the first time-step.

##### Equal Cells on Large Grid

We have built an understanding of how the two infection rate choices interact when there are two cells of equal population. Now consider the impact of the infection rate choice if all cells on a large grid are equally populated, as imagined in Figure 10(Right). How does the chosen infection rate formula interact with a spatial grid now?

Figure 11 shows simulations for the larger homogeneous grid across a range of *s*_*d*_ values. Figure 12 visualizes the relative infection rates generated at the first time step under different modeling choices. All simulations under 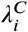 appear to have a slower invasion rate into neighboring cells than 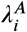,even when all other modeling choices are held constant, but this effect is only obvious for *s*_*d*_ values below a certain threshold as we see in Figure 11. For 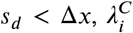predicts infection spreading slower, and therefore reaching a lower epidemic peak later than what is predicted under 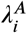.

**Figure 12:**
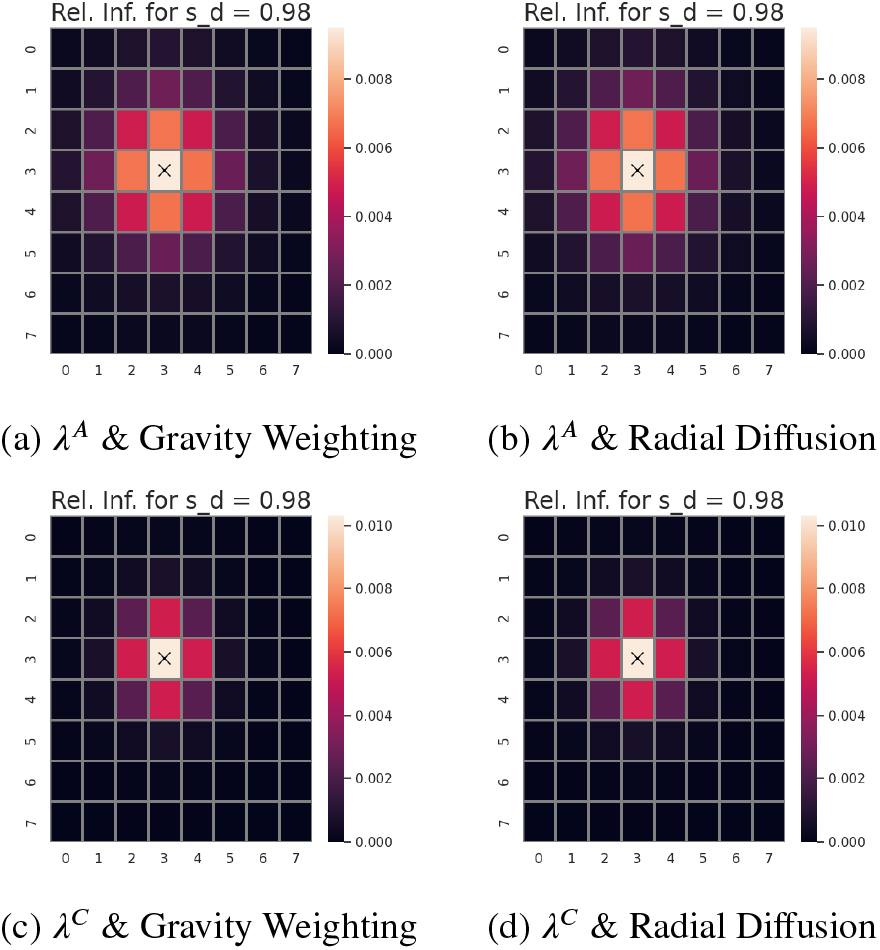
Comparison of the relative infection rate at the initial time step given a homogeneous population (Figure 10). Each row represents a different infection rate 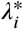 and interaction kernel *κ*^*T*^ (Tabletop Kernel) combination. This simulation used *β* = 0.5, *μ* = 0.1, and *s*_*d*_ = 1km.

#### Heterogeneous Populations

Now we aim to better understand how the infection rates interact with heterogeneous populations on a grid. As before, we start with a 2 cell example, before expanding to a larger 8 by 8 cell grid to review how these interactions scale.

##### 2 Differing Cells

What happens if the two cells now have different populations? We explicitly explore a 2 cell model as seen in Figure 13 to see how the infection rates adapt.

**Figure 13:**
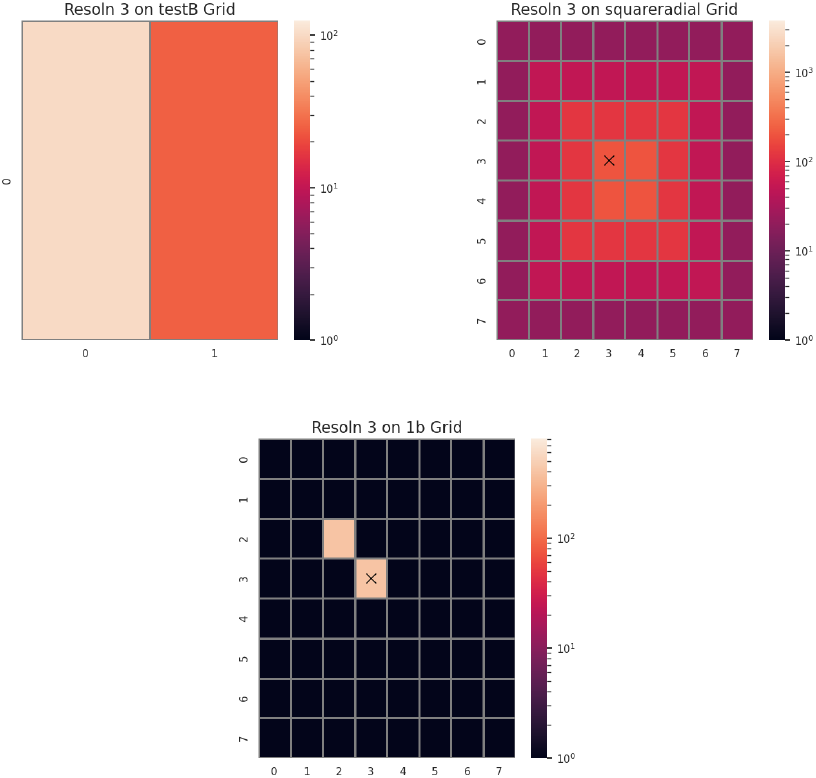
Heterogeneous population grids where Δ*x* = 1km. Top Left: Visualization of 2 cell or 2 population grid, such that cell 1 has 1/4 the population of cell 0. Top Right: Square radial population grid, designed to be an approximation of a densely populated center surrounded by less populated rings. Bottom: Sparse population of two centrally located but adjacent populated cells on an empty grid.

Averaged Infection Rate Analysis: Under 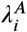 the *S*_(*i,t*+1)_ equation can be explicitly expressed as the following:

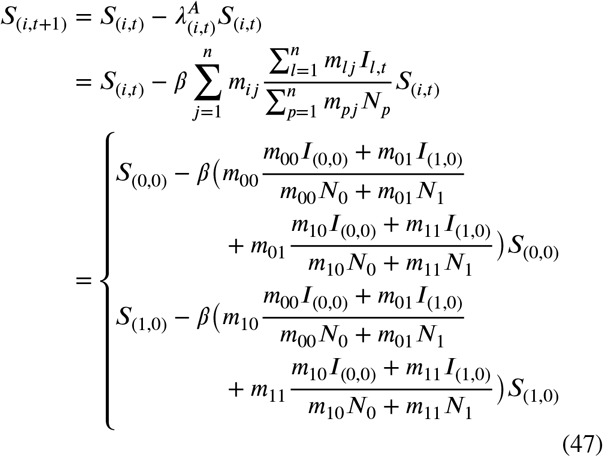

For large *N*_1_ = *N*_0_/4 and a single infection in cell 0 (that is, *I*_(0,0)_ = 1), *s*_*d*_ = 1km, these equations above become under 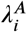:

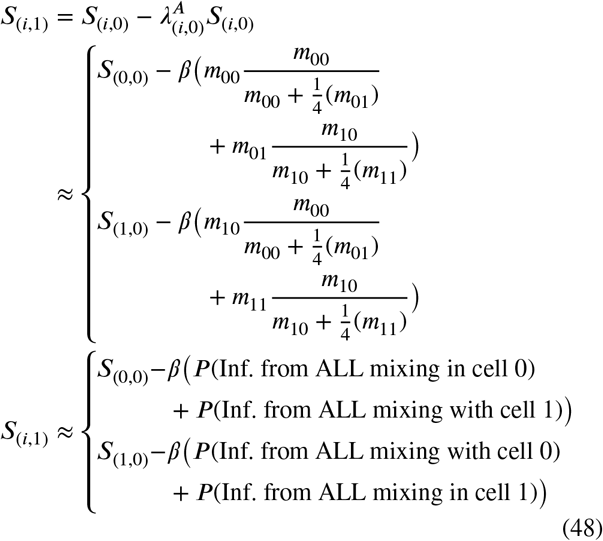

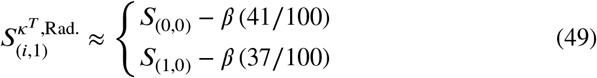

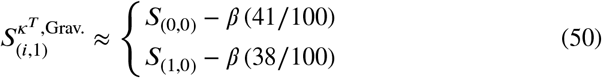

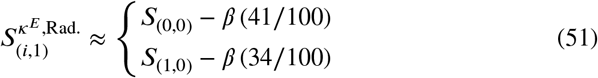

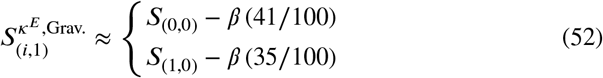

Using *κ*^*E*^, we can approximate the relative infection rates all cells under radial and gravity weighting normalization to be Equation (49) and Equation (50). Note that the relative ratio between cell 0 and cell 1 is almost maintained as it was for Equation (41), especially when compared to the dramatic shift in this same relative ratio for the Exponential interaction kernel *κ*^*E*^.

Using *κ*^*E*^, they instead approximate to be Equation (51) and Equation (52) respectively. Here, the relative ratio between cell 0 to cell 1 ratio has shifted dramatically from what was observed for without neighbors in Equation (42).

Per Capita Infection Rate Analysis: Repeating this exercise for 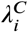, the *S*_(*i,t*+1)_ equation can be explicitly expressed as the following:

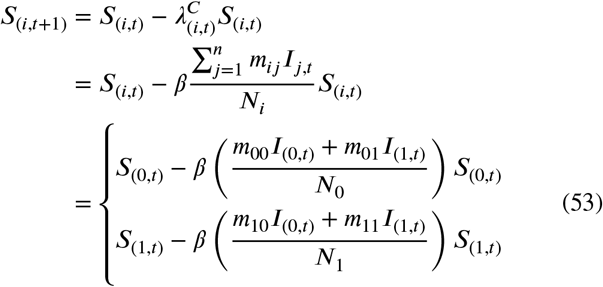

And for large *N*_0_ = *N*_1_ and a single infection in cell 1 (that is, *I*_(1,0)_ = 1), *s*_*d*_ = 1km, under 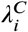:

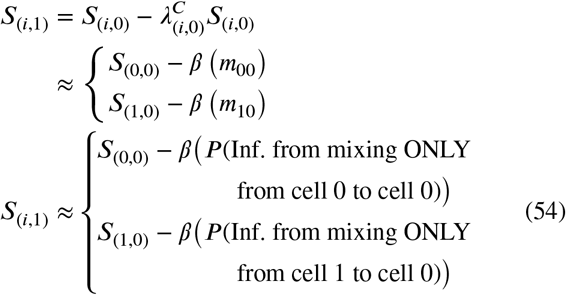

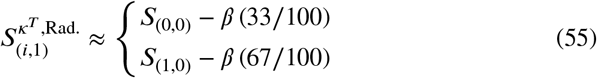

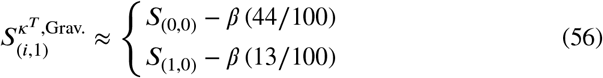

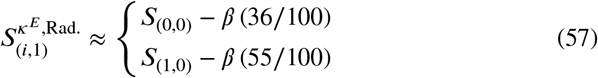

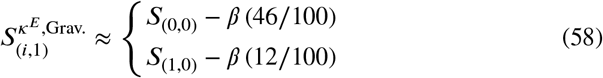

As observed with 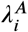, *κ*^*T*^ best maintains the ratio between cell 0 to 1 to what was seen for the equally populated neighbor case. The relative infection rate ratio between cell 0 and 1 is still larger than what was observed under the Averaged Infection rate 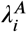 for Equation (45).

More interesting is that the ratio itself flips under radial normalization to weight the *lower* populated cell with the higher relative probability of infection, no matter the underlying interaction kernel (see Equation (49) and Equation (57)).

##### Differing Cells on Large Grid

Next, consider the impact of the infection rate choice if the entire grid is populated, but cells can have differing population densities, as imagined in Figure 13 (Top Right). How does the chosen infection rate formula interact with a spatial grid and multiple *varying* populations?

Figure 15 shows the relative infection rates at the first time step under varying model choices. Immediately apparent is the lack of pattern in the infection spread under 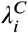 compared to the plots for 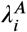.Relatively high infection rates can be observed for border cells of 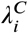 under the exponential interaction kernel (see Figure 15(d)). These border cells have relatively low populations, which under 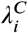 appears to inflate the realized infection rate. Since the impact of the infection rate choice have a more pronounced impact on the per-cell infection dynamics, sample of the per-cell prevalence rates are shown in Figure 14.

**Figure 14:**
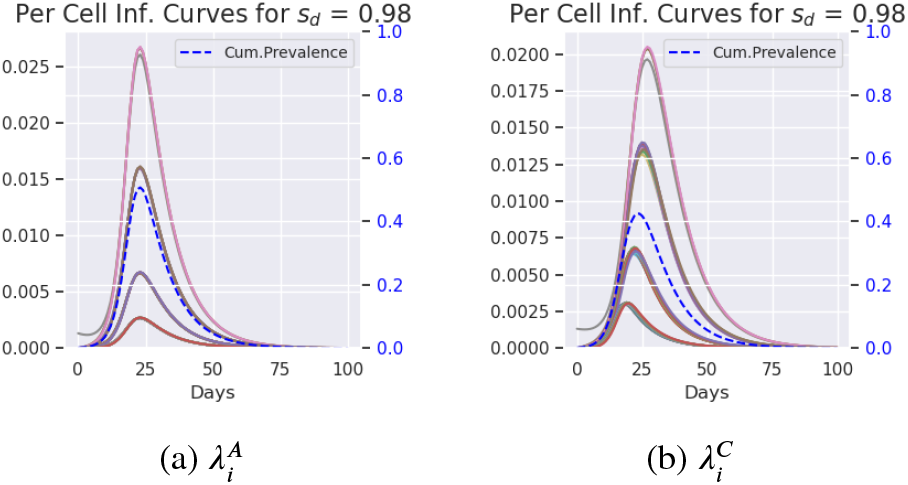
Comparison of per cell infection curves for *κ*^*T*^ (Tabletop Kernel) and gravity weighting on a square radial population (Figure 13 Top Right). The left y-axis is *Per-cell Prevalence*, while the right y-axis is *Cumulative Prevalence*. This simulation used *β* = 0.5, and *μ* = 0.1. The epidemic peak is reached faster and is greater under 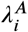 than under 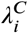 for all *s*_*d*_ choices in the figures.

**Figure 15:**
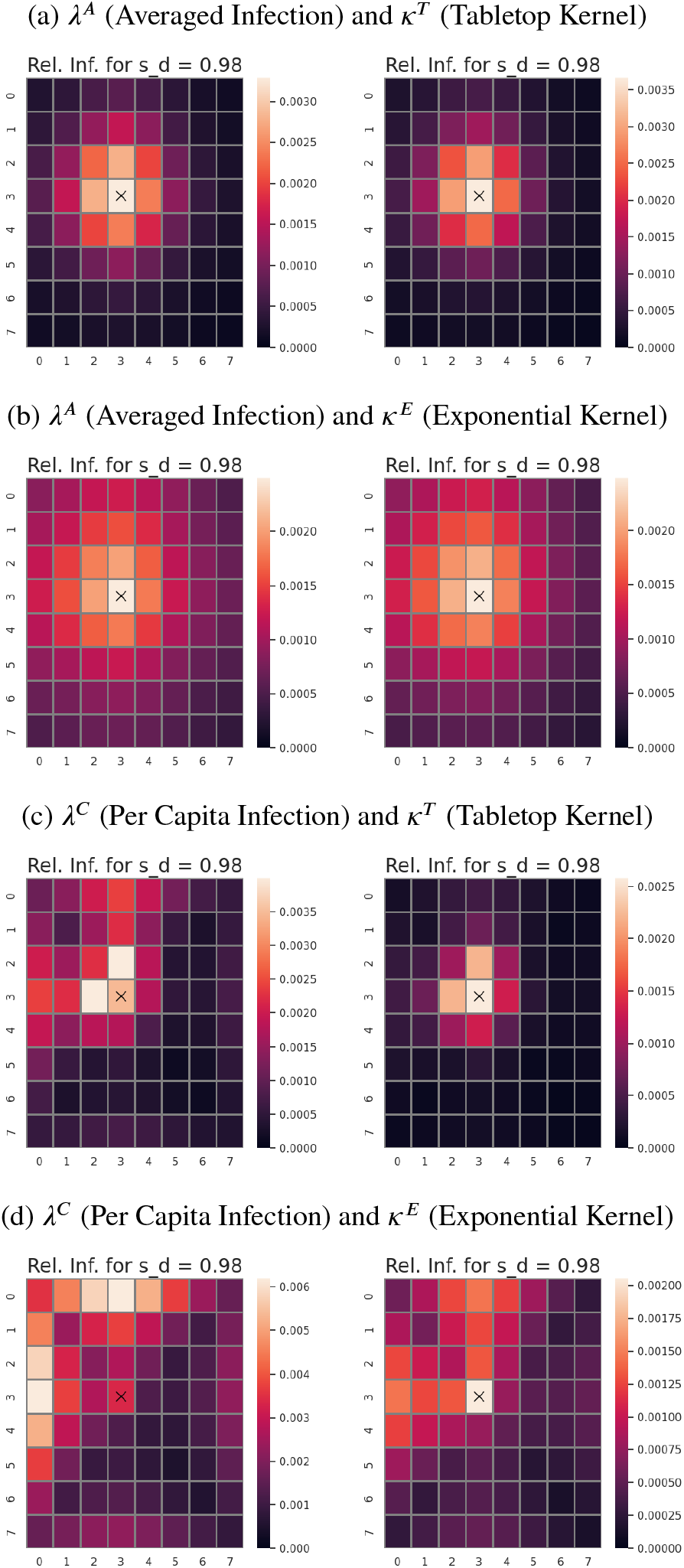
Comparison of the relative infection rate given square radial population as seen in Figure 13. Each row represents a different infection rate 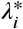 and interaction kernel κ^*^ combination. The left column shows results under gravity weighting normalization while the right column shows the results under radial diffusion normalization. The base transmission rate for this simulation was *β* = 0.5.

##### Sparse Population

Finally, consider the impact of the infection rate choice if empty cells are present on the grid, as seen in Figure 13 (Bottom). How does the chosen infection rate formula interact with empty cells on the grid?

As can be seen in Figure 2(a) (b) and (c), as the characteristic distance *s*_*d*_ increases under 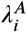,the total fraction infected increases.

The reverse happens under 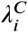 and radial normalization (Figure 2(d)). In this case, as *s*_*d*_ *decreases* the total fraction infected increased.

Further, multiple peaks can now be observed for low *s*_*d*_ values. This phenomena has been noted in the literature (Mills and Riley, 2014). Mills and Riley (2014) note the existence number of epidemic peaks is potentially over-estimated.

## Notes

### Competing Interest Statement

The authors have declared no competing interest.

## References

Verifying Scenario Planning for Geographically Diverse Populations Banos, A., Corson, N., Gaudou, B., Laperrière, V., Rey Coyrehourcq, S., 2015. The importance of being hybrid for spatial epidemic models: a multi-scale approach. Systems 3, 309–329.

Bardosh, K., Leach, M., Wilkinson, A., 2016. The limits of rapid response: Ebola and structural violence in West Africa, in: One Health. Routledge, pp. 74–94.

Bausch, D.G., Rojek, A., 2016. West Africa 2013: re-examining Ebola. Emerging Infections 10, 1–37.

Cao, W., Zhu, J., Wang, X., Tong, X., Tian, Y., Dai, H., Ma, Z., 2022. Optimizing spatio-temporal allocation of the COVID-19 vaccine under different epidemiological landscapes. Frontiers in Public Health 10, 921855.

Czocher, J., Stillman, G., Brown, J., 2018. Verification and Validation: What Do We Mean? Mathematics Education Research Group of Australasia.

Dixon, L., Sun, H., Roberts, H., 2019. African swine fever. Antiviral research 165, 34–41.

Ferdousi, T., Moon, S.A., Self, A., Scoglio, C., 2019. Generation of swine movement network and analysis of efficient mitigation strategies for African swine fever virus. PloS one 14, e0225785.

Jones, S.G., Kulldorff, M., 2012. Influence of Spatial Resolution on Space-Time Disease Cluster Detection. PLOS ONE 7, e48036. doi:10.1371/journal.pone.0048036.

Koopman, J., 2004. Modeling infection transmission. Annu. Rev. Public Health 25, 303–326.

LaBute, M.X., McMahon, B.H., Brown, M., Manore, C., Fair, J.M., 2014. A flexible spatial framework for modeling spread of pathogens in animals with biosurveillance and disease control applications. Int. J. Geo-Information 3, 638–661.

Lai, S., Ruktanonchai, N.W., Carioli, A., Ruktanonchai, C.W., Floyd, J.R., Prosper, O., Zhang, C., Du, X., Yang, W., Tatem, A.J., 2021. Assessing the effect of global travel and contact restrictions on mitigating the COVID-19 pandemic. Engineering 7, 914–923.

Lee, E.K., Li, Z.L., Liu, Y.K., LeDuc, J., 2021. Strategies for vaccine prioritization and mass dispensing. Vaccines 9, 506.

Long, E.F., Nohdurft, E., Spinler, S., 2018. Spatial resource allocation for emerging epidemics: A comparison of greedy, myopic, and dynamic policies. Manufacturing & Service Operations Management 20, 181–198.

Ma, J., Earn, D.J., 2006. Generality of the final size formula for an epidemic of a newly invading infectious disease. Bulletin of mathematical biology 68, 679–702.

Mills, H.L., Riley, S., 2014. The Spatial Resolution of Epidemic Peaks. PLoS Computational Biology 10, e1003561. doi:10.1371/journal.pcbi.1003561.

Mourant, J.R., Fenimore, P.W., Manore, C.A., McMahon, B.H., 2018. Decision Support for Mitigation of Livestock Disease: Rinderpest as a Case Study. Frontiers in Veterinary Science 5. doi:10.3389/fvets.2018.00182.

Pearson, M., Colombo, F., Cecchini, M., Scarpetta, S., 2020. Flattening the COVID-19 peak: Containment and mitigation policies. The Organisation for Economic Co-operation and Development (Issue March). https://read.oecd-ilibrary.org/view.

Pujante-Otalora, L., Canovas-Segura, B., Campos, M., Juarez, J.M., 2023. The use of networks in spatial and temporal computational models for outbreak spread in epidemiology: A systematic review. Journal of Biomedical Informatics, 104422.

Thacker, B.H., Doebling, S.W., Hemez, F.M., Anderson, M.C., Pepin, J.E., Rodriguez, E.A., 2004. Concepts of model verification and validation. Los Alamos National Lab., Los Alamos, NM (US).

Tildesley, M.J., Brand, S., Brooks Pollock, E., Bradbury, N., Werkman, M., Keeling, M.J., 2019. The role of movement restrictions in limiting the economic impact of livestock infections. Nature sustainability 2, 834– 840.

Tsai, Y.S., Huang, C.Y., Wen, T.H., Sun, C.T., Yen, M.Y., 2011. Integrating epidemic dynamics with daily commuting networks: building a multilayer framework to assess influenza A (H1N1) intervention policies. Simulation 87, 385–405.

van den Driessche, P., Watmough, J., 2008. Further Notes on the Basic Reproduction Number, in: Morel, J.M., Takens, F., Teissier, B., Brauer, F., van den Driessche, P., Wu, J. (Eds.), Mathematical Epidemiology. Springer Berlin Heidelberg, Berlin, Heidelberg. volume 1945, pp. 159– 178. doi:10.1007/978-3-540-78911-6_6.

Vergu, E., Busson, H., Ezanno, P., 2010. Impact of the infection period distribution on the epidemic spread in a metapopulation model. PloS one 5, e9371.

Wardle, J., Bhatia, S., Kraemer, M.U., Nouvellet, P., Cori, A., 2023. Gaps in mobility data and implications for modelling epidemic spread: a scoping review and simulation study. Epidemics 42, 100666.

Wu, J.T., Riley, S., Leung, G.M., 2007. Spatial considerations for the allocation of pre-pandemic influenza vaccination in the United States. Proceedings of the Royal Society B: Biological Sciences 274, 2811–2817.

Xiao, X., Newman, C., Buesching, C.D., Macdonald, D.W., Zhou, Z.M., 2021. Animal sales from Wuhan wet markets immediately prior to the COVID-19 pandemic. Scientific reports 11, 1–7.

Yashima, K., Sasaki, A., 2014. Epidemic process over the commute network in a metropolitan area. PloS one 9, e98518.

Yu, H., Wu, J.T., Cowling, B.J., Liao, Q., Fang, V.J., Zhou, S., Wu, P., Zhou, H., Lau, E.H., Guo, D., et al., 2014. Effect of closure of live poultry markets on poultry-to-person transmission of avian influenza A H7N9 virus: an ecological study. The Lancet 383, 541–548.

Zhou, Y., Xu, R., Hu, D., Yue, Y., Li, Q., Xia, J., 2020. Effects of human mobility restrictions on the spread of COVID-19 in Shenzhen, China: a modelling study using mobile phone data. The Lancet Digital Health 2, e417–e424.

